# The Genetic Determinants and Genomic Consequences of Non-Leukemogenic Somatic Point Mutations

**DOI:** 10.1101/2024.08.22.24312319

**Authors:** Joshua S. Weinstock, Sharjeel A. Chaudhry, Maria Ioannou, Maria Viskadourou, Paula Reventun, Yasminka A. Jakubek, L. Alexander Liggett, Cecelia Laurie, Jai G. Broome, Alyna Khan, Kent D. Taylor, Xiuqing Guo, Patricia A. Peyser, Eric Boerwinkle, Nathalie Chami, Eimear E. Kenny, Ruth J. Loos, Bruce M. Psaty, Tracy P. Russell, Jennifer A. Brody, Jeong H. Yun, Michael H. Cho, Ramachandran S. Vasan, Sharon L. Kardia, Jennifer A. Smith, Laura M. Raffield, Aurelian Bidulescu, Emily O’Brien, Mariza de Andrade, Jerome I. Rotter, Stephen S. Rich, Russell P. Tracy, Yii Der Ida Chen, C. Charles Gu, Chao A. Hsiung, Charles Kooperberg, Bernhard Haring, Rami Nassir, Rasika Mathias, Alex Reiner, Vijay Sankaran, Charles J. Lowenstein, Thomas W. Blackwell, Goncalo R. Abecasis, Albert V. Smith, Hyun M. Kang, Pradeep Natarajan, Siddhartha Jaiswal, Alexander Bick, Wendy S. Post, Paul Scheet, Paul Auer, Theodoros Karantanos, Alexis Battle, Marios Arvanitis

## Abstract

Clonal hematopoiesis (CH) is defined by the expansion of a lineage of genetically identical cells in blood. Genetic lesions that confer a fitness advantage, such as point mutations or mosaic chromosomal alterations (mCAs) in genes associated with hematologic malignancy, are frequent mediators of CH. However, recent analyses of both single cell-derived colonies of hematopoietic cells and population sequencing cohorts have revealed CH frequently occurs in the absence of known driver genetic lesions. To characterize CH without known driver genetic lesions, we used 51,399 deeply sequenced whole genomes from the NHLBI TOPMed sequencing initiative to perform simultaneous germline and somatic mutation analyses among individuals without leukemogenic point mutations (LPM), which we term CH-LPMneg. We quantified CH by estimating the total mutation burden. Because estimating somatic mutation burden without a paired-tissue sample is challenging, we developed a novel statistical method, the Genomic and Epigenomic informed Mutation (GEM) rate, that uses external genomic and epigenomic data sources to distinguish artifactual signals from true somatic mutations. We performed a genome-wide association study of GEM to discover the germline determinants of CH-LPMneg. After fine-mapping and variant-to-gene analyses, we identified seven genes associated with CH-LPMneg (*TCL1A, TERT, SMC4, NRIP1, PRDM16*, *MSRA*, *SCARB1*), and one locus associated with a sex-associated mutation pathway (*SRGAP2C)*. We performed a secondary analysis excluding individuals with mCAs, finding that the genetic architecture was largely unaffected by their inclusion. Functional analyses of *SMC4* and *NRIP1* implicated altered HSC self-renewal and proliferation as the primary mediator of mutation burden in blood. We then performed comprehensive multi-tissue transcriptomic analyses, finding that the expression levels of 404 genes are associated with GEM. Finally, we performed phenotypic association meta-analyses across four cohorts, finding that GEM is associated with increased white blood cell count and increased risk for incident peripheral artery disease, but is not significantly associated with incident stroke or coronary disease events. Overall, we develop GEM for quantifying mutation burden from WGS without a paired-tissue sample and use GEM to discover the genetic, genomic, and phenotypic correlates of CH-LPMneg.

## Introduction

As we age, our cells accumulate mutations. The vast majority of these mutations are inconsequential because they do not alter cell fitness. However, a small proportion of these mutations, termed drivers, can cause expansions of cell lineages they reside in. Recently, the age-related acquisition of leukemogenic point mutations (LPM) in whole blood, termed clonal hematopoiesis of indeterminate potential (CHIP), has been described as a prevalent aging-related phenomenon^1–4^. CHIP has previously been associated with increased risk for hematologic malignancy, cardiovascular disease, and increased mortality^4–6^. However, CHIP is a highly specific clonal phenomena defined as the presence of a driver mutation in 74 genes that have previously been associated with hematologic malignancy^7^, which is a small proportion of the entire spectrum of somatic variation. Non-CHIP somatic variation in blood, which we term CH-LPMneg, has previously been shown to be a prevalent phenomenon, including mosaic chromosomal alterations (mCAs)^8–10^, X-chromosome inactivation skewing^11^, and even clonal expansions without known drivers^11,12^. However, the germline determinants and clinical consequences of CH-LPMneg remain uncharacterized.

We previously used the count of high variant-allele fraction (VAF) passenger mutations in 5,071 CHIP carriers in TOPMed to identify the genetic determinants of clonal expansion, an approach termed PACER^13^, which uses age at blood-draw and passenger burden to infer the date at which a driver mutation was acquired. However, PACER is only defined for donors with a single driver mutation. Here, we seek to extend our inference of the sample level mutation burden for donors that may have no known driver point mutations. Non-CHIP clonal phenomena, which we refer to as CH-LPMneg, including mCAs, LOY, X-chromosome inactivation, and clonal expansions without known drivers have been previously associated with infection^14^, hematologic malginancy^11^, and heart failure^15^, highlighting the value of quantifying CH-LPMneg.

The accurate detection of somatic mutations in CHIP non-carriers from a single whole-blood draw is likely to be more challenging than the identification in CHIP carriers because the passenger count no longer tracks the history of a single expanded clone. We reasoned that improved estimation of the somatic mutation rate would facilitate more accurate passenger burden inference in this more challenging setting. Previous reports have identified chromatin state as among the primary determinants of mutation rate. Indeed, Shuster-Bockler and Lehner^16^ reported that variation in chromatin organization explains 55% of the variation in mutation rate. Thus, external epigenomic annotations are informative for estimating the likelihood that a candidate somatic variant call is a true mutation by altering the prior probability that a given variant call is accurate.

We and others have previously reported that CHIP and other clonal phenomena have germline genetic determinants^9,11,13,17–20^. CHIP has been previously associated with two primary pathways influencing HSC self-renewal and DNA damage pathways. Similarly, GWAS of mCAs have similarly reported associations with HSC self-renewal and DNA repair related loci^21–23^. A recent analysis that defined CH based on the dichotomization of low-VAF mutation burden, termed barcode-CH^24^, observed several hits linked to both mCAs and CHIP. These analyses have demonstrated that germline variation is associated with acquired genetic variation and have demonstrated the utility of such analyses for discovering critical regulators of clonal expansion rate, including *TCL1A*.

Here, using 51,399 donors from NHLBI TOPMed consortium^25^ without CHIP, we developed a mutation burden estimator, the Genomic and Epigenomic Mutation (GEM) rate (Figure 1). We then used this estimator as a phenotype to discover the genetic determinants of CH-LPMneg. In contrast to barcode-CH, this is a continuous phenotype that excludes individuals with CHIP mutations. This analysis revealed multiple novel loci, including the previously underappreciated role of *NRIP1*, a highly conserved transcriptional co-activator, in modulating mutation burden. We performed a sensitivity analysis, excluding all individuals with mCAs, finding that the bulk of our genetic discovery was unchanged, suggesting that CH-LPMneg signals are not merely mediated through mCAs. Fine-mapping of the *TCL1A* locus revealed a more complex cis-regulatory architecture than observed in CHIP. Functional characterization of *SMC4* and *NRIP1* with colony forming unit (CFU) assays revealed convergent effects on HSC self-renewal as the primary mechanism of these genes. Sex-stratified analyses of GEM revealed that the *TRIM59-KPNA4-SMC4* locus, which has previously been associated with CHIP and MPNs^17,26,27^ is a female-specific signal, and a novel male-specific signal near *MSRA*. Principal component analysis of mutation burden revealed a sex specific mutation pathway. GWAS of this sex-specific mutation pathway identified a novel locus near *SRGA2PC*. Through transcriptomic analyses of blood and non-blood tissues, we identified the genomic consequences of elevated mutation burden in whole blood, which include the systematic down-regulation of the interferon-alpha pathway across hematopoietic lineages. Finally, we show that GEM is useful for predicting risk of incident peripheral artery disease, and associates with altered blood cell indices. Overall, we demonstrate a novel computational approach for quantifying mutation burden, which enabled the discovery of novel genetic determinants of CH-LPMneg.

**Figure 1:**
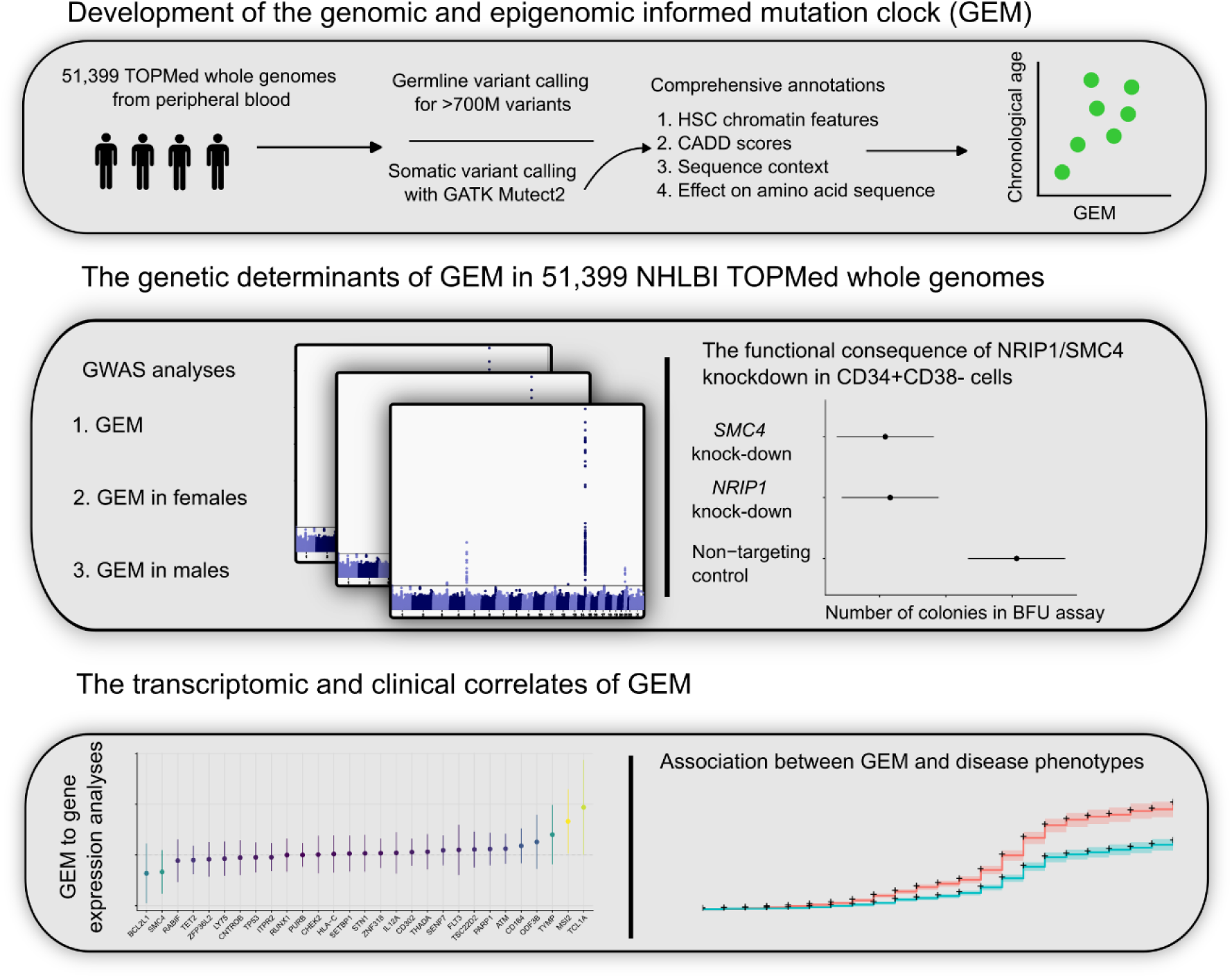
Study design schematic, describing the development of GEM, the use of GEM to discover the genetic determinants of mutation burden in blood, and the use of GEM to identify the transcriptomic and clinical correlates of mutation burden in blood.

## Results

Using 51,399 WGS samples from NHLBI TOPMed (Supplementary Tables 1-2), we first called candidate somatic variants using Mutect2 as previously described^17^. We then performed stringent filtering, including filtering known germline variants and likely sequencing artifacts (Methods). As distinguishing somatic variants from germline variants in single-tissue variant calling is challenging, we then took careful measures to determine the optimal alt-allele threshold. We observed that excluding variants with higher alt-alleles substantially improved the association of the burden of such mutations with chronological age, suggesting that a stricter alt-allele threshold than we previously applied in the PACER pipeline is useful for excluding germline variation.

### Genomic and Epigenomic Annotations Inform Mutation Rate

Next, as chromatin state is among the primary mediators of mutation rate^16^, we sought to determine the association between mutation burden and several genomic and epigenomic annotations. We used chromHMM^28^ annotations in CD34+ cells from the Roadmap Epigenomic^29^ resource as a measure of chromatin state in HSCs, which previous analyses have reported as the causal cell type in clonal phenomena^27^. We calculated the mutation burden stratified by chromatin annotation and examined the association with age, reasoning that the strength of association between mutation burden and chronological age would reflect the proportion of artifactual mutations. We observed that mutations in quiescent chromatin (Figure 2A) are much more strongly associated with age than mutations in transcriptionally active chromatin, recapitulating the role of chromatin in modifying mutation rate.

**Figure 2:**
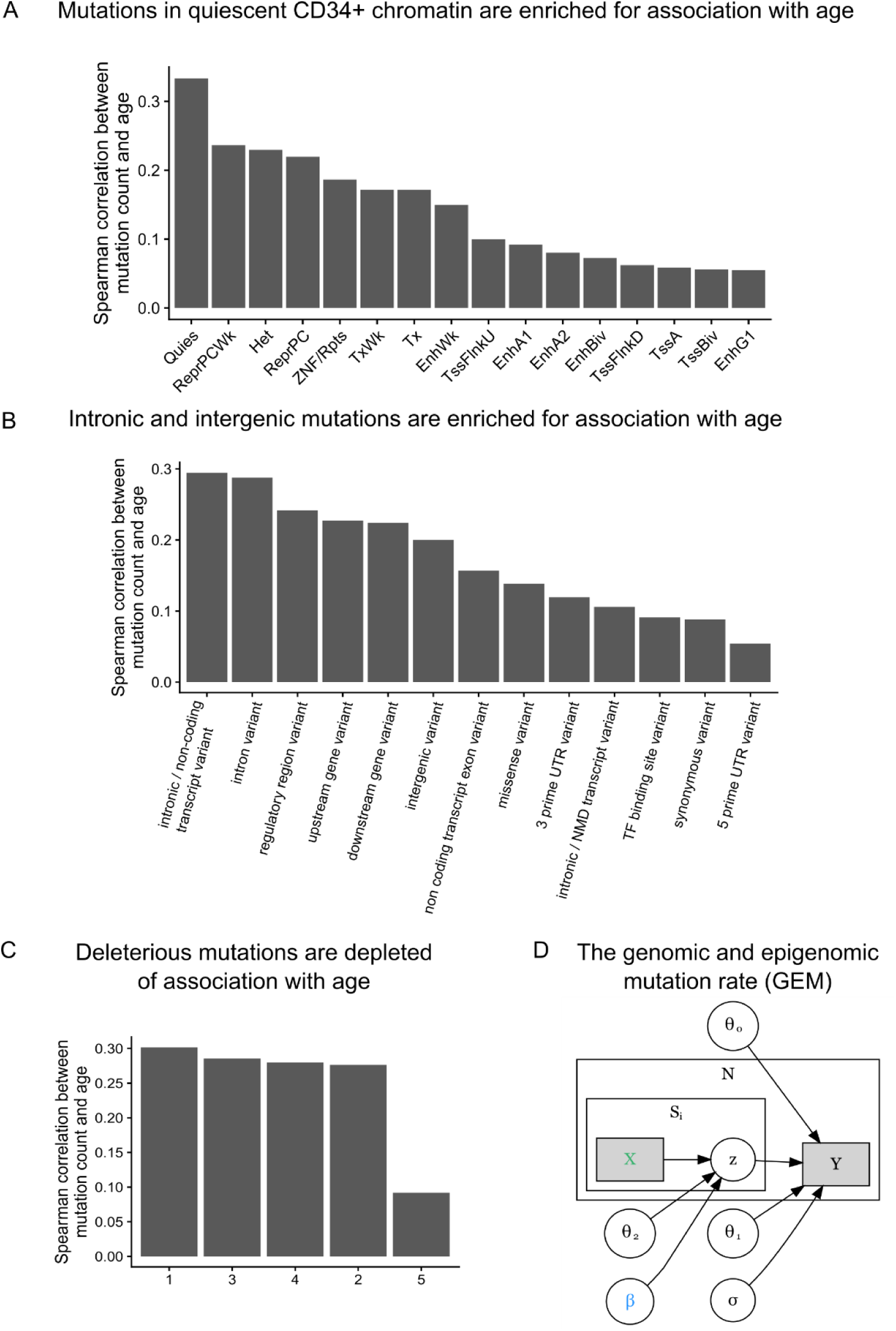
Development of GEM | A, The Spearman correlation between mutation burden and chronological age stratified by chromHMM annotations in CD34+ cells. B, The Spearman correlation between mutation burden and chronological age stratified by functional consequence as annotated by the variant effect predictor (VEP). C, The Spearman correlation between mutation burden and chronological age, stratified by quintiles of CADD scores. D, Plate annotation for the GEM statistical model. *θ*_0_and are intercepts; *θ*_1_reflects the association between log2 transformed value of ∑*z*_*ij*_and chronological age *Y*_*i*_; *z*_*ij*_ denotes the probability that the *jt*ℎ mutation in the *it*ℎ individual is a true somatic mutation. X is a matrix of annotations.

As mutations that are functional and are not mutated at the stem cell level undergo extensive negative selection^30,31^, we then asked whether mutation burden stratified by functional consequence on protein coding sequence modified the association with chronological age. We observed that mutation burden of missense and UTR variants was much more weakly associated with chronological age than mutation burden from intronic and intergenic mutations (Figure 2B), lending credence to our hypothesis that functional consequence is informative for refining mutation burden estimates. We performed a similar analysis with stratified CADD scores, finding that the mutations in the highest quintile of CADD scores were the most weakly associated with age (Figure 2C), again suggesting that deleterious mutations are depleted of association with age and likely enriched for false positives. We performed an analyses stratified by both CADD quantile and chromHMM annotation, finding that the two were not redundant (Extended Data Figure 1). Collectively, these analyses suggest that variant deleteriousness and chromatin annotations are useful for the construction of a mutation derived molecular clock.

We then developed a weakly-supervised probabilistic graphical modeling approach that incorporates genomic and epigenomic annotations to distinguish somatic mutations from artifacts, a method we term GEM (genomic and epigenomic mutation rate). Weakly-supervised probabilistic graphical modeling approaches have been previously used to identify functional rare-variants^32^, demonstrating the utility of such approaches towards annotation of genetic variation. We first comprehensively annotated all candidate somatic variants based on their chromHMM^28^ annotations, their functional consequence, CADD^33^, their population allele frequency in TOPMed, surrounding sequence context, among others (Methods). GEM uses chronological age as an external annotation to identify which candidate somatic mutations were functional based on their annotations (Figure 2D). GEM enables the classification of candidate somatic mutations as either truly somatic or artifacts, thus increasing power in downstream analyses by facilitating the depletion of likely artifactual variants.

### The Genetic Determinants of Mutation Burden

Next, as clonal phenomena have been shown to have germline genetic determinants, we performed a GWAS with GEM as the phenotype in 51,399 carriers of diverse ancestry. We computed summary statistics using SAIGE^34^. We detected six genome-wide significant loci, including *TERT, TCL1A, TRIM59-SMC4-KPNA4, NRIP1, SCARB1, and PRMD16* (Figure 3A), and an overall h^2^_SNP_ of 9.3%. We then performed a similar analysis based on the burden of mutations in quiescent chromatin and heterochromatin, which resulted in reduced power at *TCL1A* (GEM minimum pvalue of 6 x 10^-57^ vs mutation burden minimum pvalue of 6 x 10^-50^), demonstrating the value of the GEM over simply using mutation burden stratified by chromatin context (Extended Data Figure 2). *TERT*, *TCL1A*, *TRIM59-SMC4-KPNA4* have been previously associated with CHIP^17,18,35^and barcode-CH^24^, while *NRIP1* and *PRDM16*, *TERT*, and *TCL1A* have all been associated with mCAs^21–23^ and barcode-CH^24^. *SCARB1* has not been previously reported with a related phenotype. To nominate causal SNPs and genes, we fine-mapped each locus using SuSIE^36^ and cross-referenced the credible sets with the Open Targets V2G estimates^37^ and cell-type specific enhancers-gene pairs from the activity by contact model^38^. These signals collectively highlight the convergence of germline variation influencing CHIP, mCAs, and clonal hematopoiesis without known drivers.

**Figure 3:**
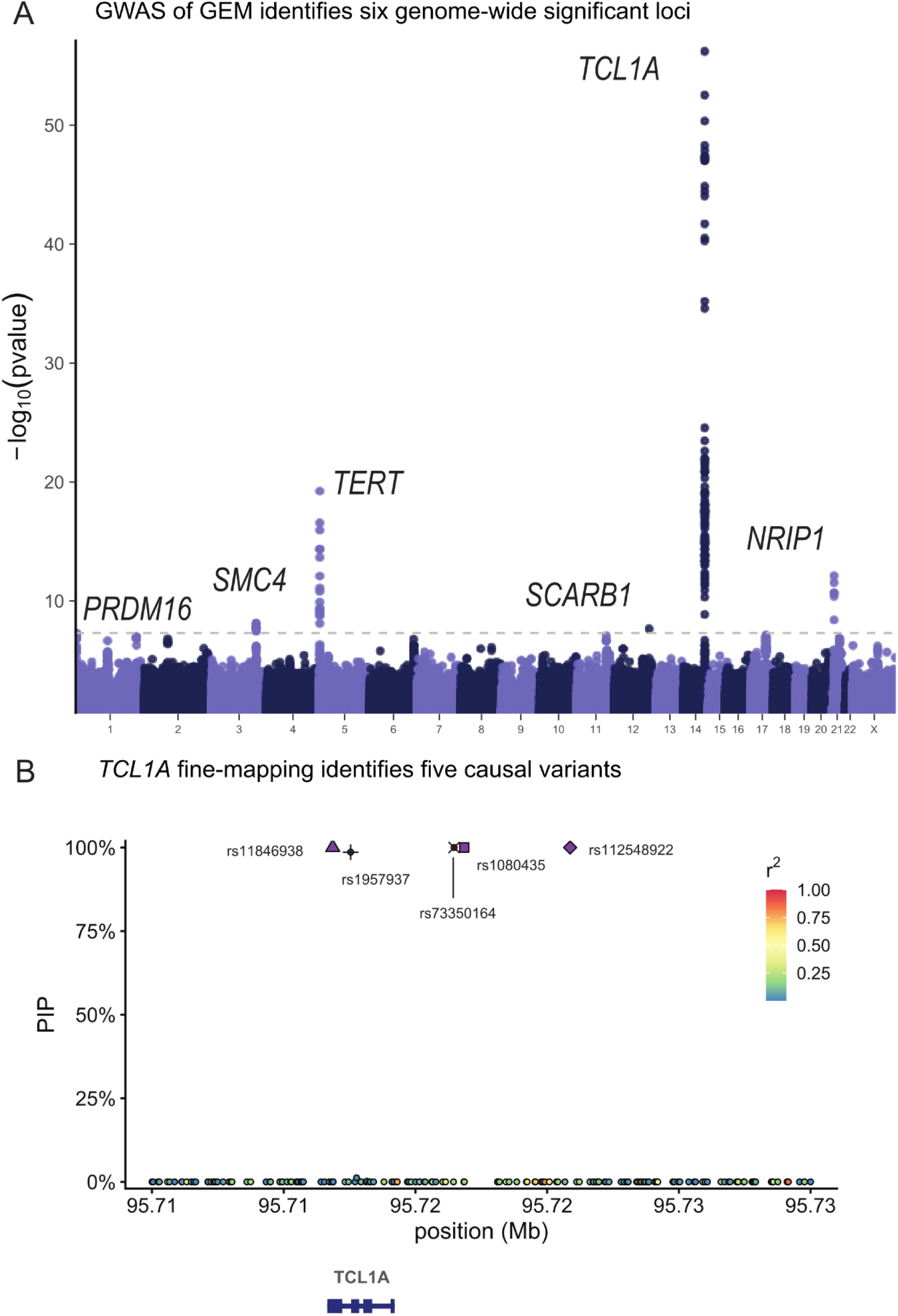
The genetic determinants of GEM. A, The GWAS of GEM. Summary statistics were estimated with SAIGE. B, Fine-mapping of the TCL1A locus. Note rs11846938 is 10bp from rs2887399. Fine-mapping was performed with SuSIE.

We then asked whether the association between these GWAS loci and GEM was mediated through an association with mCAs. Using a recently developed atlas of mCAs in TOPMed^20^, we excluded all mCAs or LOX carriers (n = 13,399) and performed another GWAS as a sensitivity analysis. After filtering to variants that were genome-wide significant in either GWAS, we observed that the effect sizes were remarkably concordant (R^2^ = 0.997, Extended Data Figure 3), suggesting that the GWAS of GEM is not merely mediated by the effect of mCAs/LOX.

The lead variant at *TCL1A* was rs2887399, which we previously discovered as among the primary mediators of clonal expansion in CHIP carriers^39^, and has been previously reported in GWAS of LOY^21,22^. Fine-mapping^36^ revealed five credible sets, each with a single causal variant (rs11846938, rs112548922, rs1080435, rs1957937, and rs73350164, all PIP > .98). V2G^37^ identified *TCL1A* as the mostly likely causal gene at each of the five causal SNPs. rs11846938 is 10bp from and in very high LD with rs2887399 (EUR R^2^ = 0.88, AFR R^2^ = 0.92), and both reside in the core promoter of *TCL1A*. Given high LD between rs2887399 and rs11846938, and the functional evidence we previously observed for the effect of rs2887399, we refer to this signal as “rs2887399/rs11846938.” rs112548922 is 9kb upstream of *TCL1A*, suggesting that cis-regulatory elements besides the core promoter are implicated in altered mutation burden. We previously described a CHIP-mutation specific mechanism, whereby mutations in *TET2/ASXL1/SF3B1* are associated with the aberrant chromatin opening of the *TCL1A* promoter in HSCs, but not *DNMT3A* mutations. This led us to hypothesize that rs2887399 is chromatin accessibility-QTL (caQTL) that is a *TET2/ASLX1/SF3B1* mutation (and possibly LOY) specific and thus the risk allele is more likely to lead to the aberrant activation of proto-oncogene *TCL1A* only when a mutation sufficient for chromatin modification at the *TCL1A* promoter has been acquired. Indeed, rs2887399 has been since reported as a caQTL^40^ in lymphoblastoid cell lines (LCL). We previously showed that *TCL1A* expression is sufficient for promoting clonal expansion and altering stress response in HSCs^39^ and is a key regulator of clonal expansion in CHIP clones. The discovery of additional casual variants at the *TCL1A* locus highlights the utility of applying GEM to samples unascertained for specific genetic lesions, providing increased power for genetic discovery and indicates that the context specific up-regulation of *TCL1A* is a broader phenomenon than previously appreciated, and likely occurs in HSCs without mCAs, LOY, or CHIP, possibly mediated through stochastic epigenetic phenomena that result in increased accessibility at cis-regulatory elements of *TCL1A*.

The lead variant at *NRIP1* is rs2229742, a common (MAF = 6%) missense variant (p.Arg448Gly) predicted to be deleterious by SIFT^41^. *NRIP1* has previously been discovered in the context of mCAs^14^ and barcode-CH^24^ but has not been discovered in CH-LPMneg. Fine-mapping of the locus identified one credible set containing three variants (rs2229742, rs2823020, rs2823025). The C allele of rs2229742 is associated with increased GEM burden (beta = 0.08 standard deviations, 72% PIP, pvalue = 7.6x10^-13^). *NRIP1* is a highly conserved (pLI^42^ = 0.99) transcription co-regulator that is highly expressed in HSCs^43^ and has been previously reported as a positive regulator of stemness in HSCs^44^. rs2229742 is strongly associated with multiple blood cell index GWAS ^45^ indicating that altered protein sequence of *NRIP1* results in altered HSC function. *NRIP1* ablation has previously been shown to extend lifespan in murine models^46^, indicating a role in aging related phenotypes, although this is reported to be likely mediated through the interaction between *NRIP1* and estrogen signaling rather than modulation of HSC function. As the C allele is associated with increased GEM, this suggests that the C allele may either increase function of the *NRIP1* product or increase the abundance of *NRIP1* through indirect mechanisms, perhaps through increased translation efficiency. To elucidate the consequences of altered amino acid sequence in *NRIP1,* we cross-referenced a recently released catalogue of *trans-*pQTLs from plasma^47^. rs2229742 is a trans-pQTL for both SDC4 (beta = 0.09, pvalue = 3.8 x 10^-13^) and PGLYPR2 (beta = 0.07, pvalue = 8.7 x 10^-14^). SDC4 is a syndecan, which are cell-surface proteins that can interact with a broad range of ligands. The mouse-genome informatics resource^48^ reported that *SDC4* ablation in mice led to several altered hematopoietic phenotypes. *PGLYRP2* is a peptidoglycan recognition protein that has been implicated in interferon regulation and innate immune response^49^.

Previous GWAS of clonal phenomena, including CHIP and MPNs have reported the *TRIM59-SMC4-KPNA4* locus, though none have conclusively identified the causal gene. Fine-mapping this locus identified a credible set containing 19 variants. The three variants with the highest PIP were rs11718121 (PIP = 9.2%), rs1451760 (PIP = 8.8%), and rs6790951 (PIP = 8.2%). V2G estimated that *SMC4* was the mostly likely causal gene for each of these three variants, and alt-alleles at the three variants were associated with increased expression of *SMC4* in eQTLGen in whole blood^50^ and increased expression of *SMC4* in lipopolysaccharide stimulated monocytes^51^. The interval spanned by the credible set contains a predicted *SMC4* enhancer in CD34+ cells by the ABC model. *SMC4* is a sub-unit of the condensin complex, which is involved in chromosome assembly and segregation during mitosis. Collectively, fine-mapping, V2G estimates, and the ABC model nominate *SMC4* as the mostly likely causal gene in the locus, highlighting the role of *SMC* related proteins in modulated mutation burden.

Fine-mapping of the *SCARB1* locus identified one credible set with a single SNP, rs11057853, a common (MAF = 46%) variant intronic to *SCARB1*. V2G estimated that *SCARB1* was the most likely causal gene for rs11057853, supported by both its presence within the *SCARB1* gene body and its role as an eQTL for *SCARB1* in blood^50^. The C allele was associated with reduced GEM (beta = -0.03 GEM standard deviations) and reduced expression of *SCARB1* (beta = -0.23, pvalue = 5.5 x 10^-175^), suggesting that *SCARB1* may be protective against CH-LPMneg. *SCARB1* is a receptor for HDL and rare variant burden tests of *SCARB1* in UK Biobank have identified several associations with lipid traits^52^. Previous reports have described a possible role for *SCARB1* in mediating the metabolic adaptation of long-term HSCs using murine models^53^.

We then asked whether rare variants are associated with GEM. We performed a genome-wide RVAS using STAAR^54^, including all missense and loss of function (LOF) variants within protein-coding genes. We identified 33 and 18 hits at pvalue thresholds of 5 x 10^-6^ and 5 x 10^-7^ (Supplementary table 3). The strongest hit was *CELF2* (pvalue = 3.4 x 10^-28^), where coding variants were associated with a higher GEM value among carriers (mean of 0.38, 95% CI: [0.36, 0.40]) than non-carriers (mean of 0.00, 95% CI: [-0.01, 0.01]). *CELF2* is a highly constrained (pLI^42^ = 1.00) RNA binding protein and is highly expressed in neutrophils^37^. A recent report described the role of *CELF2* as a suppressor of the AKT/PI3K signaling pathway^55^ in lung carcinoma, which is presumably the signaling pathway mediating the effect of *TCL1A*.

We performed an non-coding RVAS using SCANG^56^, a dynamic window approach for identifying sets of SNPs that associate with phenotypes. We applied to SCANG to 100kb regions flanking 1,688 genes that Open Targets^37^ has identified as previously associated with cancer (Supplementary table 4). We identified 52 and 6 hits at pvalue thresholds of 5 x 10^-6^ and 5 x 10^-7^ (Supplementary table 5). We observed a locus on chr14 flanking the MARK3 gene which was strongly associated with mutation burden (2.5 x 10^-8^). To identify the likely causal variants within this window, we then performed a joint analysis of all rare variants that were included. This analysis highlighted three variants as significantly associated including rs190231639, a rare (MAF = 0.04%) variant intronic to *COA8*. To identify the likely causal gene in this gene dense locus, we cross-referenced the V2G results from Open Targets, which nominated *COA8*, *KLC1*, *XRCC3*, and *ZFYVE21* as equivalently likely causal genes. *XRCC3* is involved in the homologous recombination repair pathway of double-stranded DNA, though we are unable to conclusively identify it as the causal gene. Collectively, we identify diverse signals among the rare variants implicating DNA repair and post-translational modifications as key molecular processes contributing to GEM variation.

### Mutation Burden Is Indirectly Regulated by the Size of the HSC Pool

We then asked whether *SMC4* and *NRIP1* contributed to mutation burden by altering HSC self-renewal. To test this hypothesis, we separately knocked down *SMC4* and *NRIP1* in CD34+ bone-marrow derived human HSCs with shRNAs and performed a colony forming unit (CFU) assay. Relative to a non-targeting shRNA control, both *SMC4* and *NRIP1* knockdown cells were more likely to lose stemness in culture (1.6% and 2.5% fewer CD34+CD38-cells, pvalues = 2.0 x 10^-3^, 1.7 x 10^-3^, Figure 4A-B) and the cells formed fewer burst-forming unity colonies (22.0 and 22.8 fewer colonies, pvalues = 2.4 x 10^-3^, 1.9 x 10^-3^, Figure 4C-D). These results are consistent with a role of both *SMC4* and *NRIP1* as positive regulators of CD34+ HSC self-renewal, and led us to propose the following mechanisms for their roles as indirect regulators of mutation burden: Either *SMC4/NRIP1* may regulate the fitness of HSC in response to noxious stimuli such as the infection inherent to the shRNA knockdown assay, or *SMC4/NRIP1* directly regulates the size of the HSC pool; a larger active HSC pool increases the likelihood that at least one HSC obtains a fitness advantage through either a genetic lesion or stochastic epigenetic phenomena, leading to a clonal expansion, which will increase the passenger count burden. Similar models have been previously been proposed in the context of myeloproliferative neoplasms^27^.

**Figure 4:**
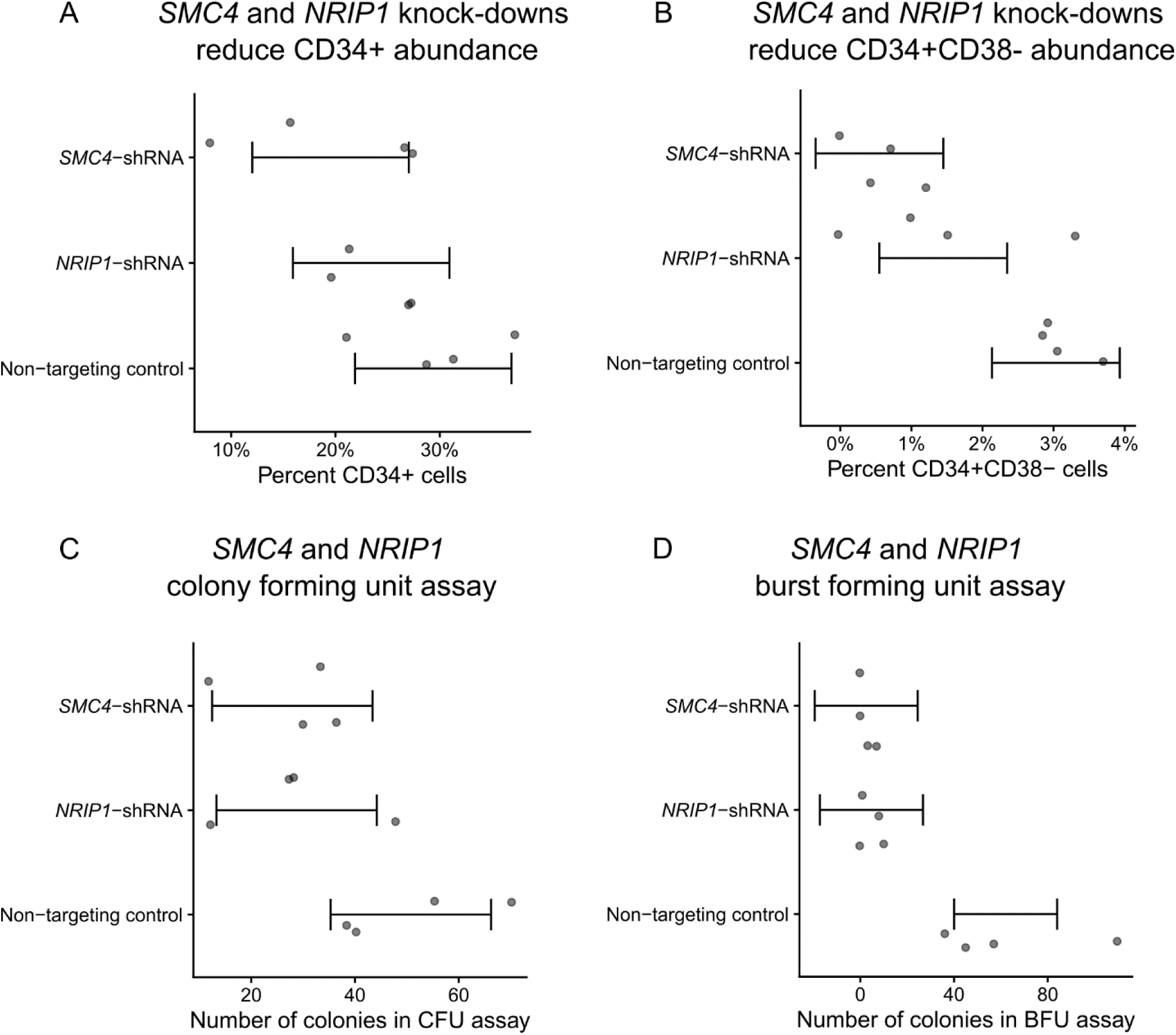
The functional consequence of SMC4 and NRIP1 on HSCs. A, SMC4 and NRIP1 were knocked-down with shRNA and the proportion of CD34+ cells was quantified with FACs. Quantities were compared referent to a non-targeting control. B, proportion of CD34+CD38-was quantified with FACS. C, Number of colonies formed in a colony-forming unit (CFU) assay. D, Number of colonies in a burst-forming unit assay.

We then sought to explore the causal relationship between HSC pool size and GEM through simulation using a stochastic process that describes realistic HSC population (Methods). We simulated several HSCs which acquire passenger mutations at a constant rate per cell through a Poisson point process. We simulated the size of individual HSC clone populations using a Poisson birth-death process, where a single parameter *s* governs the relative likelihood of an HSC self-renewing into two identical HSCs as opposed to dividing into two differentiated cell types. At a rate of 1 driver mutation per 10,000 HSCs per year, we simulated modest increases to *s* in each HSC to model modest increases in cell fitness that some clones may acquire. We stratified these simulations across varying initial sizes of the HSC pool, finding that larger pools were much more likely to contain at least one clone with a substantial increase in fitness (Extended Data Figure 4) and many more high-VAF passengers (Extended Data Figure 5). Importantly, the burden of high VAF passengers increased as the number of increases to *s* increased. Taken together, this model provides a formal exposition for why GEM may be associated with both the overall HSC pool and the likelihood of at least one clone obtaining a substantial increase in self-renewal capacity.

### Regional Mutation Burden and the Genetic Determinants of Sex Specific Mutation Pathways

We then sought to perform more granular analyses of mutation burden, examining heterogeneity by sex and position in the genome. As the importance of mutation burden may vary based on genome position, we estimated the mutation burden in 49 non-overlapping intervals approximately 5 x 10^7^ bases in length. We then estimated the association between age and mutation count stratified by interval, indicating heterogeneous associations across the intervals (Extended Data Figure 6). To characterize the underlying structure, we then performed PCA on these mutation counts.

We observed that PC1 was strongly associated with overall mutation burden and explained 60% of the variance, revealing a single general factor associated with mutation burden genome-wide. We observed that the loadings of PC2 were enriched for mutations appearing chromosome-X, suggesting a sex-specific mutation pathway. We observed that PC2 is significantly associated with genotype inferred sex (R2 = 7.3%, pvalue < 2.2 x 10^-16^), highlighting a sex-specific contribution to somatic variation in whole blood.

We then asked whether this sex-specific mutation factor had distinct germline determinants from GEM. We observed that a single locus on chromosome 1 near *SRGAP2C* was associated with PC2. The lead variant at this locus is rs61804016, a common variant 62kb away from the transcription start sites of *SRGAP2C* that has been previously reported as an eQTL in whole blood for *SRGAP2C*, *NBPF8*, *NBPF26*, *PFN1P2*, and *SRGAP2C*. However, in monocytes and T-cells, rs61804016 is only an eQTL for *SRGAP2C*^57^. The eQTL associations and proximity to the TSS of *SRGAP2C* suggest that *SRGAP2C* is the most likely causal gene in the locus. *SRGAP2C* is a GTPase activating protein that is expressed in hematopoietic progenitor cells^43^. We then cross-referenced phewas^58^ results in UK Biobank^59^ and Finngenn ^60^. The C allele of rs61804016 is nominally associated with increased risk for breast cancer (odds-ratios of 1.08, 1.07, pvalues of 1.2 x 10^-5^, 3.2 x 10^-6^), further supporting the sex-specific nature of PC2. *SRGAP2C* is on chromosome 1, suggesting sex-specific regulation of an autosomal gene in the genesis of sex-specific mutation burden. No SNP near *SRGAP2C* was associated at genome-wide significance with the GEM phenotype. These analyses highlight the value of subtyping in mutation burden estimation by revealing sex-specific factors.

Given findings of sex-specific mutation pathways, we then performed sex-stratified GWAS of GEM (Figure 5), which revealed two sex specific signals. At the *SMC4* locus, rs11718121 was associated with increased mutation count in females (beta = 0.043, pvalue = 5.1 x 10^-9^) but much more weakly associated in males (beta = 0.019, pvalue= 0.038). At a locus not identified in the standard analysis, near *MSRA*, rs117344298 was associated with decreased mutation count in males (beta = -0.17, pvalue = 2.4 x 10^-8^), but unassociated in females (beta = -0.013, pvalue = 0.58). Other signals were largely shared between males and females. Cross-referencing of sex-biased eQTLs reported in GTEx^61^ found that *MSRA* had nominally significant sex-biased eQTLs in tibial nerve tissue and Brain cortex, but not in whole blood. *SMC4* did not have sex-biased eQTLs, which may be the result of limited power to detect such effects.

**Figure 5:**
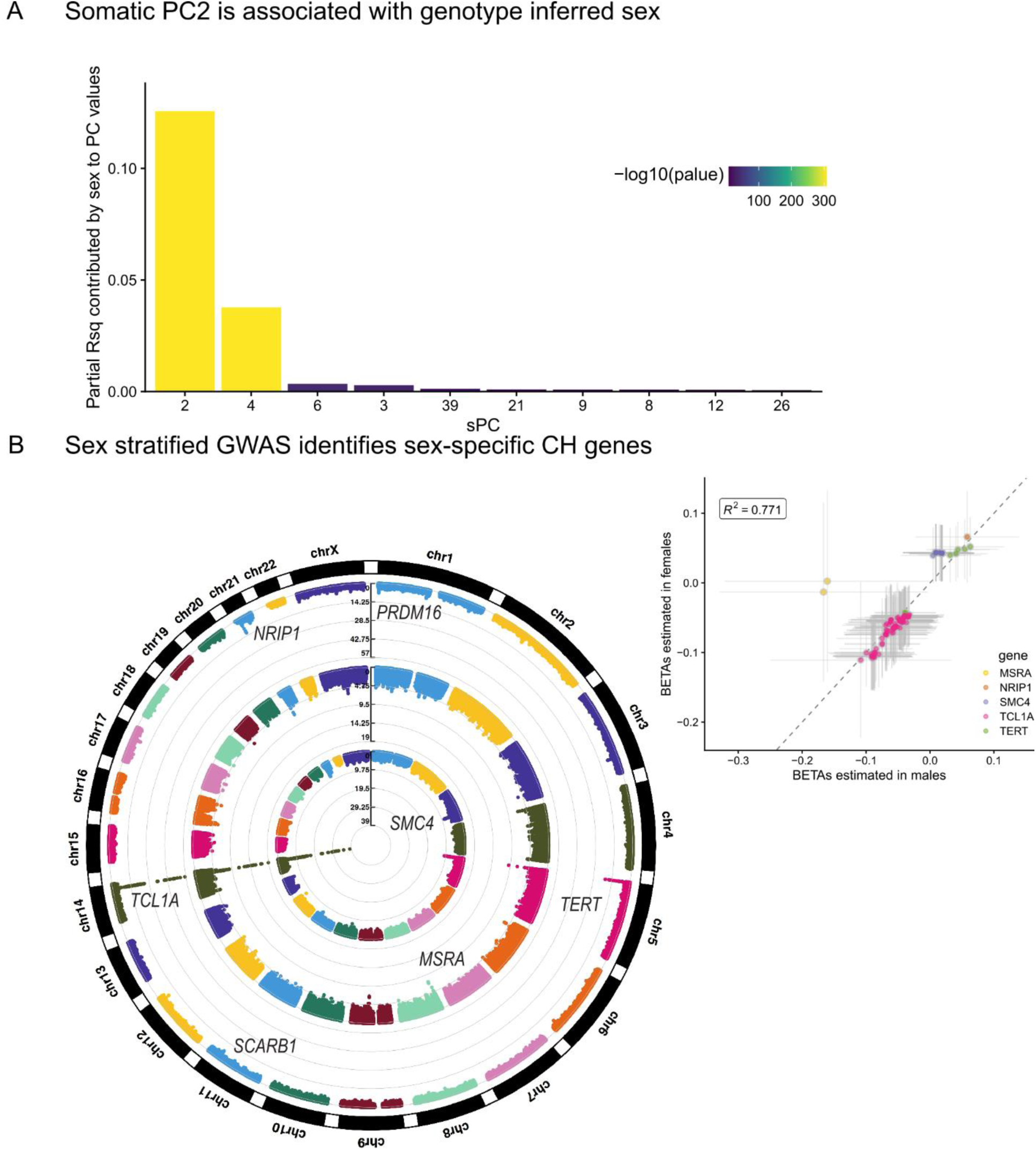
The sex specific genetic determinants of mutation burden. A, Regressions were performed for each quantile-transformed somatic principal component (sPC) on study and sex as covariates. The partial variance explained by sex is displayed on the y-axis. B, Circular Manhattan plot. The outer-most ring is the GWAS of GEM on all individuals, the middle ring is the GWAS of GEM on males, and the inner-most ring is the GWAS of GEM in females. Inset, a scatter plot of the two sex-specific GWAS plotting all SNPs with pvalues < 1 x 10^-8^ in either GWAS. Asymptotic confidence intervals are plotted with a width corresponding to genome-wide significance.

### The Transcriptomic Correlates of High Mutation Burden

Next, we asked whether GEM associated with altered gene expression across five tissue types available in TOPMed, including whole blood, PBMC, monocytes, T cells, and nasal epithelial cells. We performed a search for GEM-gene associations by regressing the inverse normal transformed expression values of each gene (n = 17,741) on the inverse normalized GEM estimates, including age, genotype inferred sex, 15 genotype PCs, 20 expression PCs, and cohort indicators as covariates. To increase power, we then used mashr ^62^ to apply shrinkage across the 88,705 GEM-gene associations. We identified 404 GEM-gene associations at a local false sign rate (lfsr)^63^ < 0.05 (Supplementary Table 6). Within whole blood, we observed the up-regulation of *RUFY4* with increased GEM. *RUFY4* is highly expressed in dendritic cells^37^ and is involved in response to the anti-inflammatory cytokine IL-4^49^ (Figure 6A). We also observed the down-regulation of *LGSN*, which although annotated for its role in differentiation cells in the lens, is highly expressed in HSCs and was recently reported as a candidate causal gene in asthma^64^.

**Figure 6:**
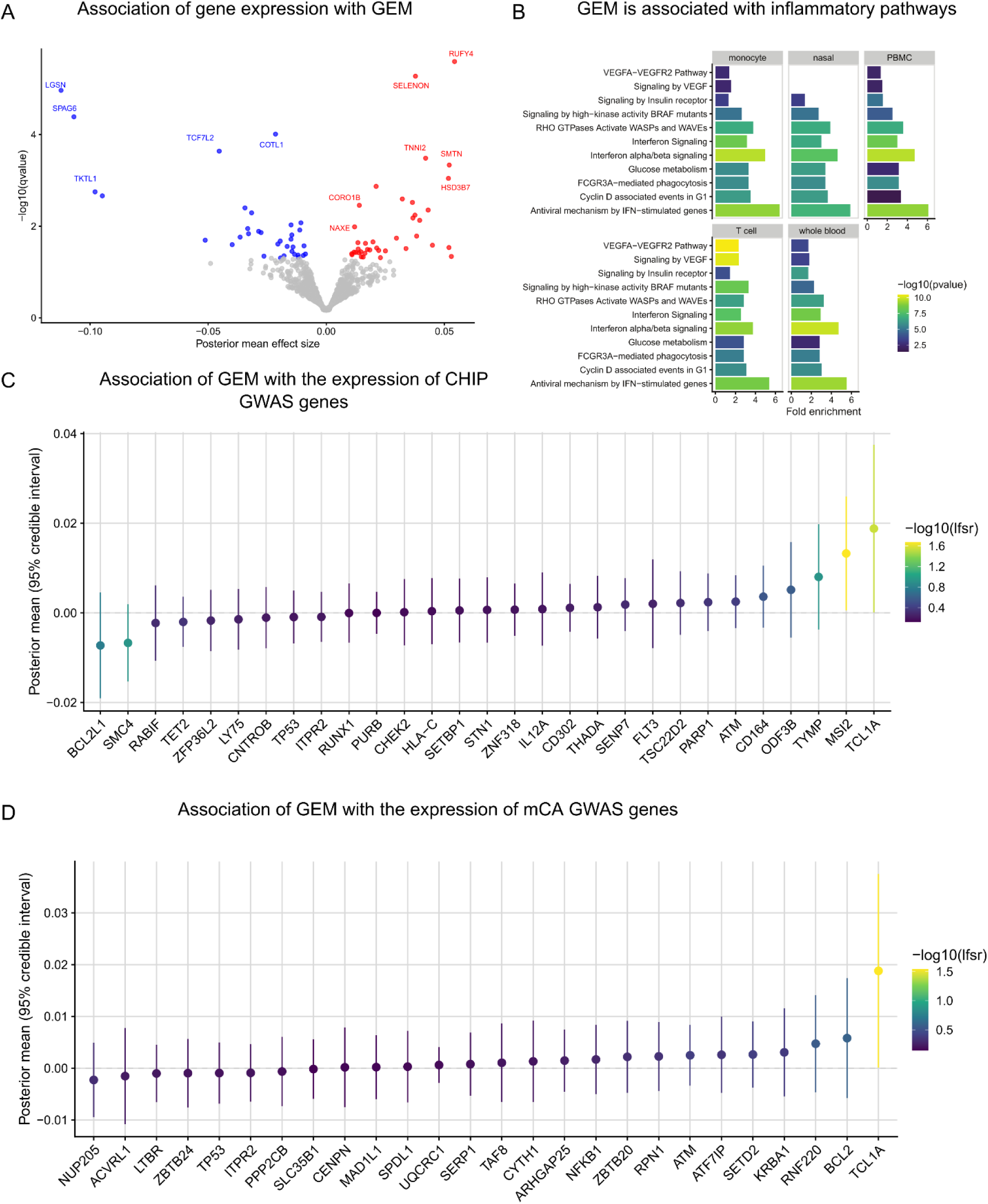
The transcriptomic correlates of GEM. A, Association analyses were performed between GEM and gene expression in whole blood, including age, sex, genotype PCs 1-5, and expression PCs 1-20 as covariates. B, Enrichment analyses were performed using pathfindR and KEGG pathways as reference. C, Association statistics among CHIP GWAS genes. D, Association statistics among mCA GWAS genes.

To characterize the gene programs associated with GEM, we performed pathway enrichment analyses using KEGG^65^ as a reference. We observed a striking down-regulation of genes involved in interferon signaling (Fig. 6B). Interferon-alpha is a cytokine with anti-proliferative properties that was previously considered as a candidate therapeutic for AML^66^. Interferon-alpha is thought to reduce clonal expansion through direct and indirect mechanisms, including inducing apoptosis and activating the adaptive immune system. We then performed a tissue specific pathway analysis among the 404 GEM-genes, which similarly identified interferon alpha/beta signaling as greater than 3x fold enriched in each of the five tissue types. We also identified enrichment of the VEGFA-VEGFR2 pathway that is largely specific to T cells (Fig. 6B). Deletion of VEGFA in CD8+ T cells has previously been shown to reduce effector function^67,68^. Collectively, these results highlight the importance of anti-proliferative cytokines to inhibiting clonal expansion and suggest that transcriptomic responses to mutation burden in disparate tissues (nasal epithelial and blood samples) are more similar than anticipated.

Next, we asked whether specific loci that either define CHIP, or have been discovered in GWAS of CHIP or mCAs (Supplementary Tables 7-8), implicate genes whose expression levels are also associated with GEM. Among loci identified in either CHIP or mCA GWAS, we observed that expression of *TCL1A* and *MSI2* are positively associated with GEM (Fig. 6C-D). The association between *TCL1A* and GEM is consistent with *TCL1A* expression as a key mechanism in modulating mutation burden and clonal expansion in whole blood. Among CHIP mutations (Supplementary Table 9), we observed that *IDH2* expression is negatively associated with GEM (Extended Data Figure 7), which corroborates its protective effects against clonal expansion. We then asked whether within blood, there were specific genes with heterogeneous effects. We found that although effect sizes generally were highly concordant, *TCL1A* had a much stronger association with GEM in T cells than in monocytes (Extended Data Figure 8), highlighting the need the tissue and cell-specific transcriptomic analyses when performing searches in blood for the transcriptomic correlates of mutation burden.

### The Clinical Correlates of GEM

Because clonal hematopoiesis phenomena have been previously associated with cardiovascular disease^4,6,15^, we asked whether GEM associates with vascular and heart disease phenotypes in TOPMed. We first asked whether GEM was associated with coronary artery disease (CAD). We restricted our analyses to those NHLBI TOPMed cohorts with harmonized longitudinal assessment of CAD events; this enabled separate analyses of the association between GEM and incident CAD phenotypes (i.e., CAD events that occurred after the blood draw from which GEM was assessed) and the association between GEM and prevalent CAD (i.e., CAD events prior to the GEM blood draw). Within four NHLBI cohorts (WHI, FHS, CHS, COPDGene), we performed a Cox-proportional hazards regression analyses for incident CAD events after excluding individuals with prevalent CAD disease, including GEM, age at baseline, smoking history, body-mass index (BMI), sex, and germline genotype PCs as covariates. We observed that GEM was not associated with incident CAD events (meta-analysis hazard ratio: 1.00, 95% CI: [0.97, 1.04], pvalue = 0.84, Figure 7A).

**Figure 7:**
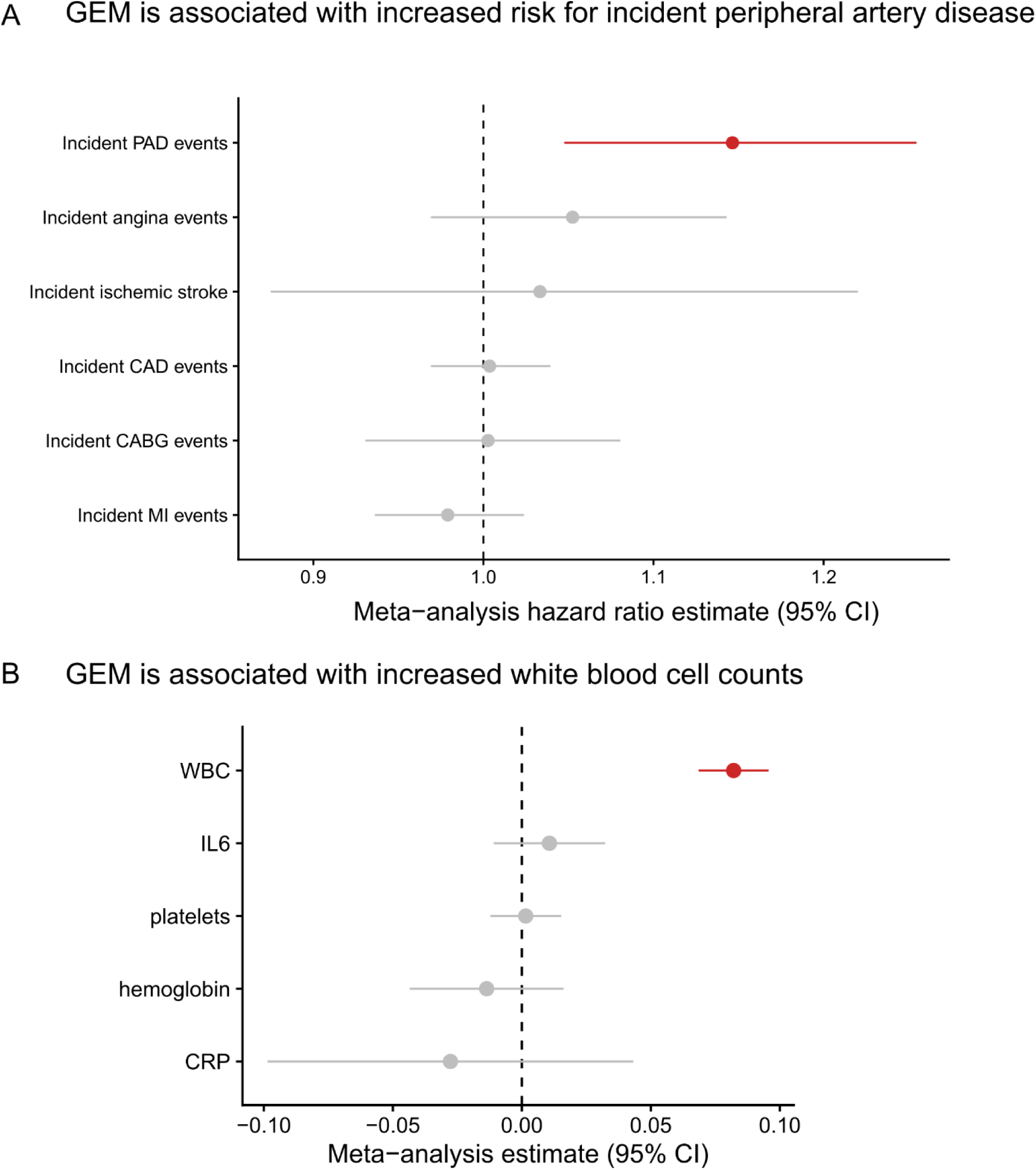
The phenotype correlates of GEM. A, Cox proportional-hazard regressions were performed, regressing incident events on a spline of age, sex, smoking status, and germline PCs. Individuals with prevalent disease were excluded. CAD = coronary artery disease, PAD = peripheral artery disease, CABG = coronary artery bypass graft, MI = myocardial infarction. CAD events were defined as at least one of an MI, CABG, angina, or angioplasty during the follow-up period. A random effects meta-analysis was performed. GEM was inverse normal transformed. Sex was excluded from the WHI regression, and smoking was excluded from the COPD regression. B, A linear regression of the inverse normal transformed biomarker, including a spline of age, sex, smoking status, and germline PCs as covariates. GEM was inverse normal transformed. Sex was excluded from the WHI regression, and smoking was excluding from the COPD regression.

We performed similar analyses with incident ischemic stroke. After meta-analyzing results from three cohorts (WHI, CHS, ARIC), we observed that there was substantial heterogeneity (I^2^ = 86%) in results, with a positive association observed in WHI (hazard ratio of 1.19, 95% CI: [1.12, 1.26], pvalue = 2.8 x 10^-8^) and null or negative effects observed in ARIC and CHS (Extended Data Figure 9). Meta-analysis resulted in no association between incident stroke and GEM (hazard ratio of 1.03, 95% CI: [0.88, 1.22]), Figure 7A). Given the differences in the distribution of sex across the three cohorts, we then performed a female only analysis in CHS and ARIC, finding no evidence for a sex-specific effect after meta-analyzing with WHI (hazard ratio of 1.05, 95% CI: [0.87, 1.26], Extended Data Figure 10). We then asked whether GEM is associated with incident peripheral artery disease (PAD). After meta-analysis, we observed that GEM was associated with increased risk (hazard ratio 1.15, 95% CI: [1.05, 1.26], pvalue = 3 x 10^-3^, Figure 7A) for incident PAD events.

Because evidence within the clonal hematopoiesis literature (Heyde et al.^69^) suggests that prior CAD events are causal contributors to the up-regulation of HSC proliferation, we then asked whether prior CAD was associated with GEM. In a meta-analysis across five NHLBI cohorts (WHI, FHS, CHS, COPD, ARIC), we observed that prior CAD was suggestively associated with a modest increase in GEM (effect of prior CAD on standardized GEM: 0.12, 95% CI: [-0.03, 0.28], pvalue = 0.13, Extended Data Figure 11).

To reconcile these disparate phenotypic associations, we performed analyses between GEM and biomarkers, including complete blood cell counts (CBC) measurements and inflammation measurements. We observed that GEM was positively associated with increased white blood cell count after adjustment for age and smoking status at baseline (Fig. 7B). These results indicate that GEM is associated with altered hematopoiesis. In contrast, after meta-analysis, we observed no association with CRP (Figure 7B). This suggests that the association between GEM and PAD is not mediated through systemic inflammatory markers like CRP, but we note that markers of systemic inflammation like CRP may not be sufficiently sensitive to capture the association between inflammation and HSC activity within the bone-marrow microenvironment.

## Conclusion

Using 51,399 diverse TOPMed whole genomes, we derived a semi-supervised model of mutation rate, GEM, that increases power for discovery of germline determinants of mutation burden by better distinguishing somatic mutations from sequencing artifacts. Using GEM to identify the germline determinants of CH-LPMneg, we observed the convergence of common variant loci influencing multiple types of clonal phenomena, including CHIP and mCAs, demonstrating that genetic predisposition to mutation burden is shared across several different clonal contexts. We observed that altered protein sequence of *NRIP1* was strongly associated with increased mutation rate, which along with its documented role in GWAS of blood cell indices, collectively implicate *NRIP1* as an important regulator of aberrant hematopoiesis. We also observed that *TCL1A*, which we previously identified as a critical moderator of clonal expansion in CHIP carriers, is also associated with mutation burden in samples ascertained for not having CHIP, which may reflect its contribution towards clonal expansion in other kinds of clonal phenomena. Using a sex specific mutation pathway revealed by PCA analysis, we identified a breast-cancer locus, *SRGAP2C*, that associates with GEM.

We anticipate that our approach, which identifies distinct underlying mutation pathways through PCA analysis can be extended to identify other factors that may contribute to specific mutation pathways. Overall, our approach identifies several loci that have been previously discovered in other analyses of clonal hematopoiesis phenomena, suggesting that studying clonal hematopoiesis without known drivers represents an under-appreciated model for discovering the germline determinants of mosaicism in blood. Importantly, analysis of mutation burden without a known CHIP genetic lesion greatly expands the sample size available to perform these analyses; our analysis here is an order of magnitude larger than the number of CHIP carriers discovered in our previous analyses of TOPMed analyses^13,17^.

We performed the first multi-tissue analysis of the transcriptomic consequences of mutation burden in whole blood. We observed the striking down-regulation of interferon signaling across five tissues, including four from blood and one from epithelial tissue. Collectively, this analysis highlighted the need for additional characterization of the *in-vivo* transcriptomic consequences of anti-proliferative cytokines on HSC growth. Interferon-alpha, among other cytokines with similar effects, may represent candidates for therapeutic intervention.

We observed that mutation burden in whole blood was associated with altered blood cell indices and increased risk for peripheral artery disease. However, GEM was not associated with incident CAD events. This is consistent with the observation that the association between CH and CAD is highly heterogenous across different forms of clonal phenomena. Within CHIP, the largest phenotype analysis to date^35^ reported an association (1.31 hazard ratio) with *TET2* CHIP but not *DNMT3A* CHIP. Several analyses within smaller cohorts have also reported associations between CHIP and CAD phenotypes^4,6,70^. CH mediated through mCAs have no reported association with CAD^9^, while both positive and negative reports exists regarding LOY and CAD related phenotypes^15,23^. A recent analysis that examined barcode-CH^24^, which includes several different forms of CH, reported no association between barcode-CH and CAD, while finding an association between barcode-CH and PAD, concordant with our results. These observations reflect the need to examine the associations between CH and CAD stratified by the particular genetic lesion(s), size of clone, and potentially the rate at which a clone is expanding. Indeed, recent reports^52,71^ on the plasma proteomic associates of CH have found substantial heterogeneity across different CHIP mutations, highlighting the substantial heterogeneity observed at both epidemiologic and molecular levels.

Our approach is not without limitations. The precise estimation of mutations from whole blood remains challenging. Although this approach has been shown to be promising in large cohorts in the context of epidemiologic association analyses, more sensitive sequencing assays are needed for clinical application. Additionally, the genesis of several mutations remains unclear. Although clonal expansion without known drivers clearly occurs^12^, elucidating the underlying mechanism remains an open question.

Overall, we develop a novel estimator of mutation burden that is not specific to CHIP carriers. We find that measuring mutation burden, even in individuals without known genetic lesions, is informative for aging related phenotypes. In contrast to surveillance for CHIP, which is relatively rare in individuals less than 80 years old, GEM can be used to monitor mutation burden in a larger proportion of adults. We anticipate that our approach will prove useful in non-blood tissues for the discovery of the germline basis of mutagenesis and will facilitate epidemiologic association analyses, ultimately elucidating the genesis and consequences of mutation burden.

## Extended Data Figures

**Extended Data Figure 1:**
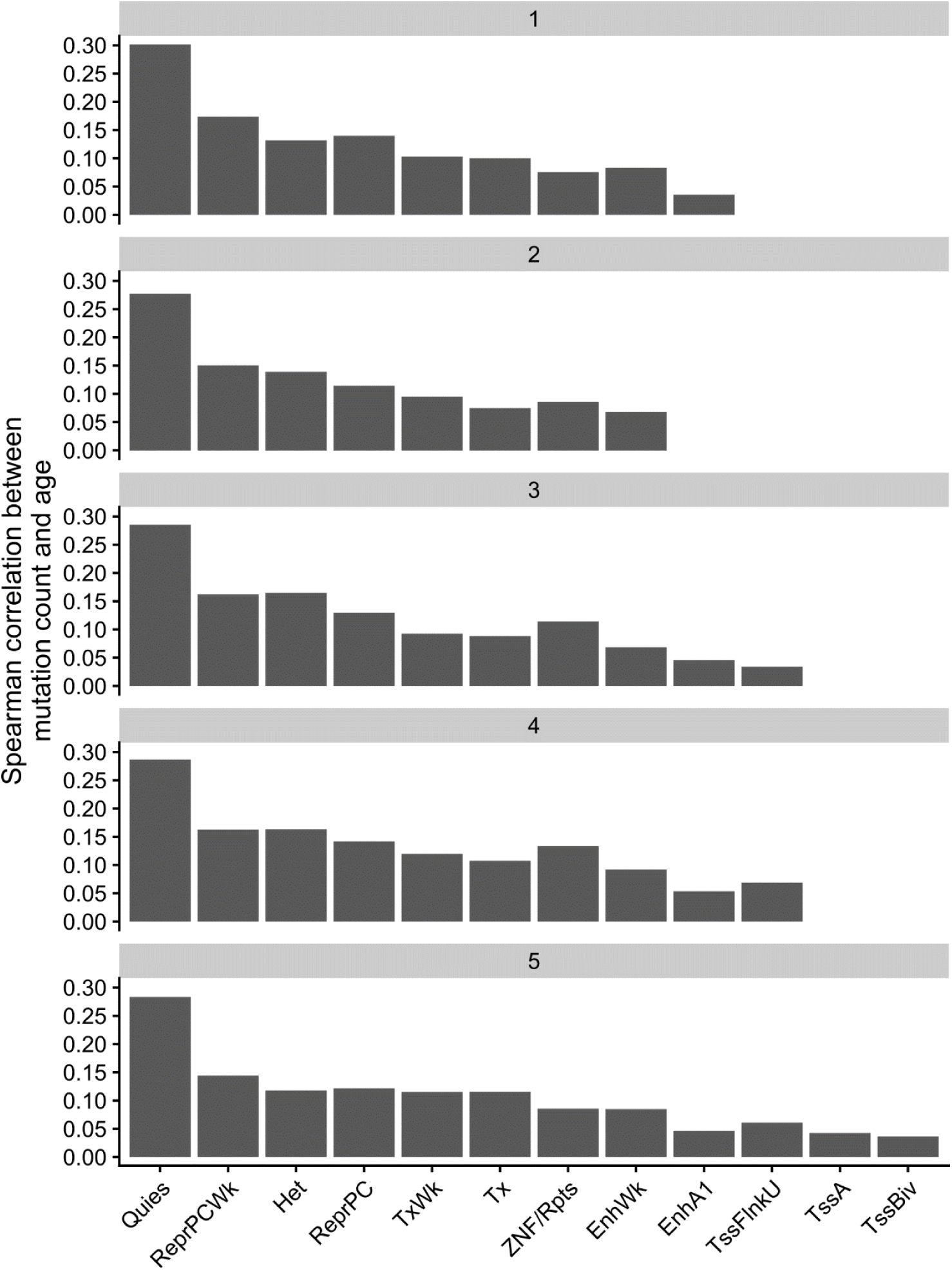
Spearman correlation between mutation burden and chronological age was calculated for each of the strata defined by chromHMM 15 state model in CD34+ cells and CADD derived quintiles. A CADD score of 5 indicates a score within the top 20% most deleterious variants.

**Extended Data Figure 2:**
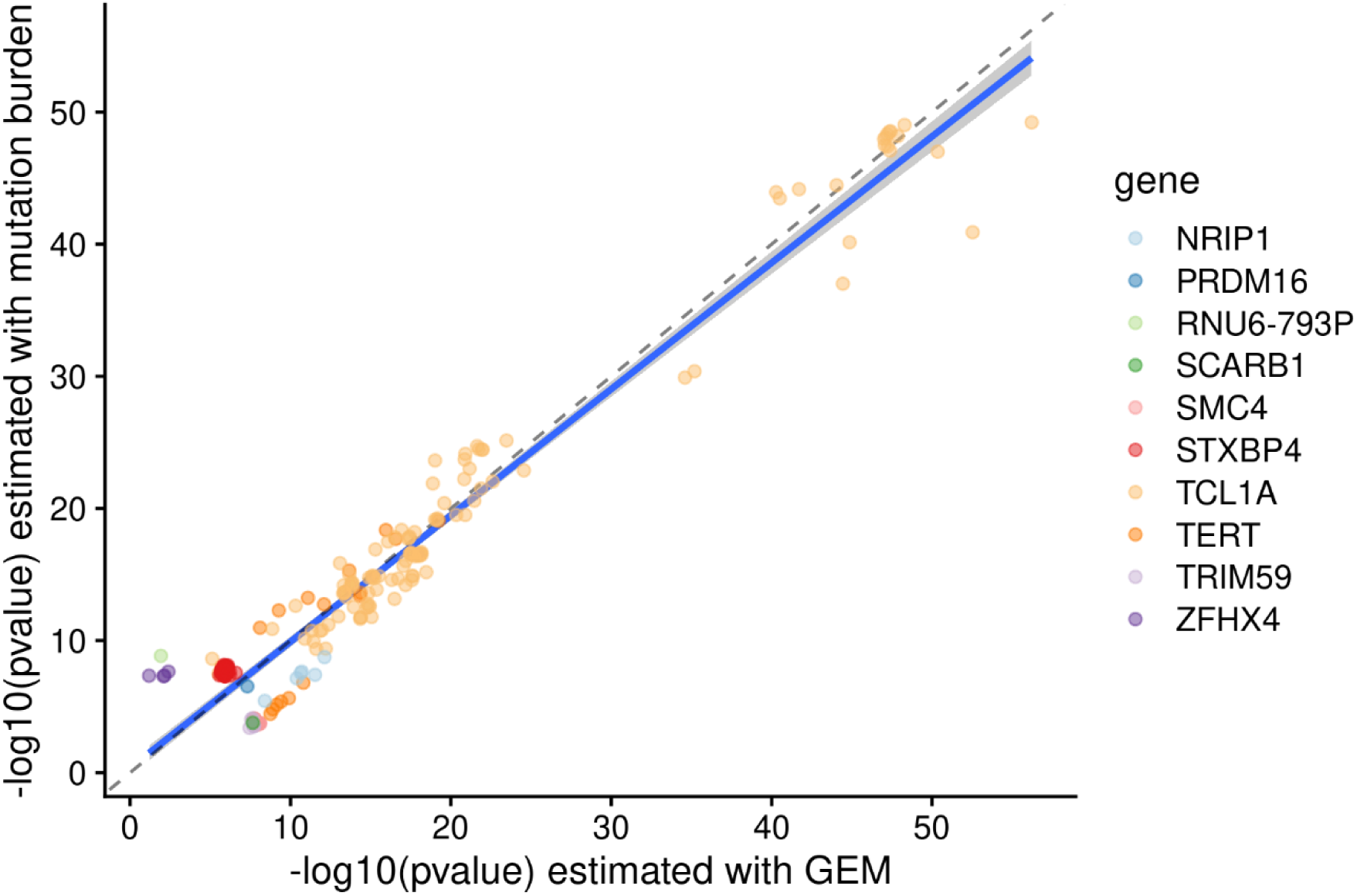
Scatter plot comparing the -log10 pvalues from GWAS where the phenotype was either GEM (x-axis) or the burden of mutations falling in either heterochromatin or quiescent chromatin in CD34+ cells. Genes are colored by the likely causal gene, which was manually curated. Variants shown have pvalue < 5 x 10^-8^ in at least one of the two GWAS.

**Extended Data Figure 3:**
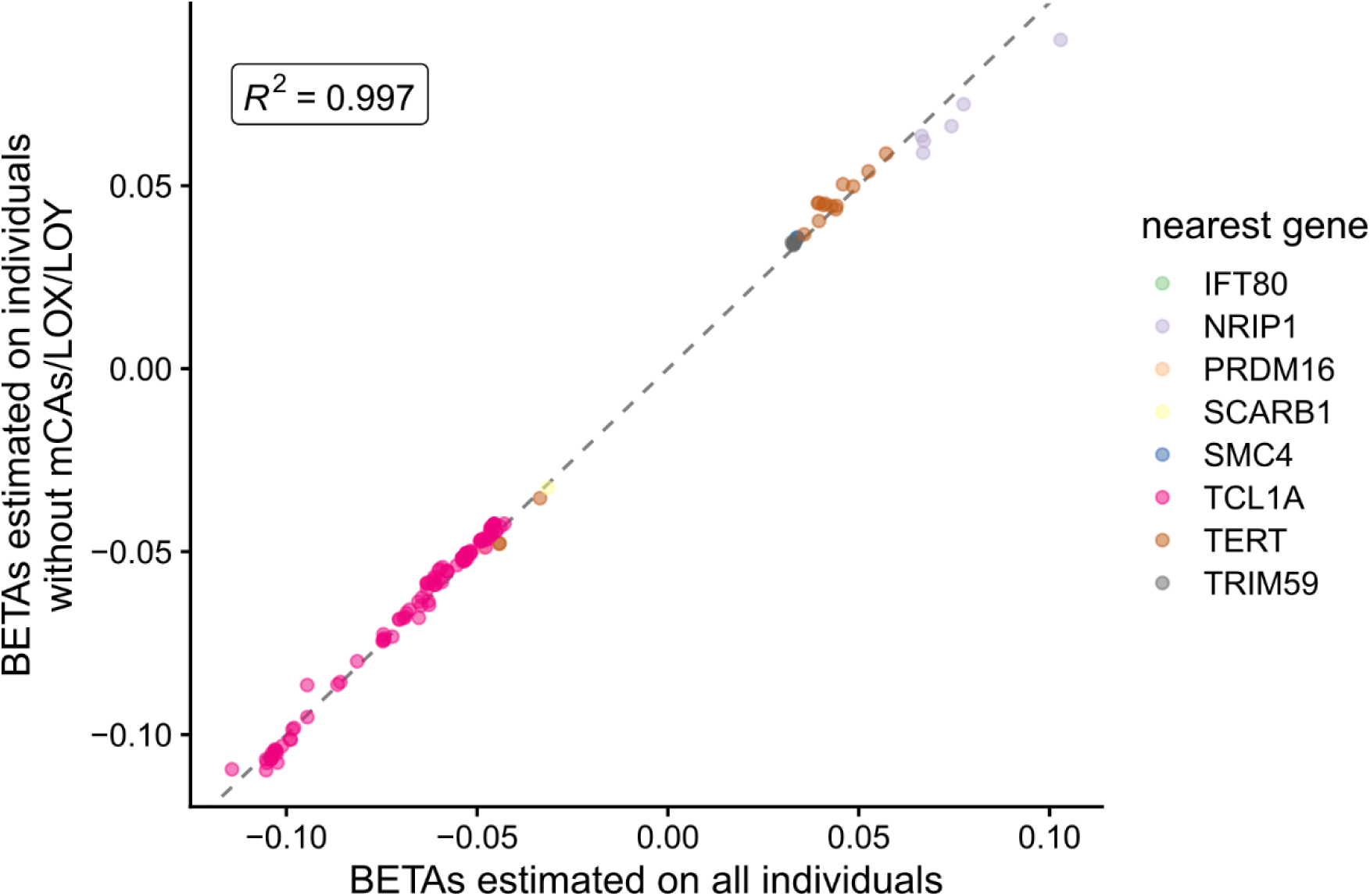
Scatter plot comparing the beta values from GWAS where the phenotype was either GEM on all individuals (x-axis, n= 51,399) or GEM on individuals that did not have an mCA (n = 38,000). Genes are colored by the likely causal gene, which was manually curated. Variants shown have pvalue < 5 x 10^-8^ in at least one of the two GWAS.

**Extended Data Figure 4:**
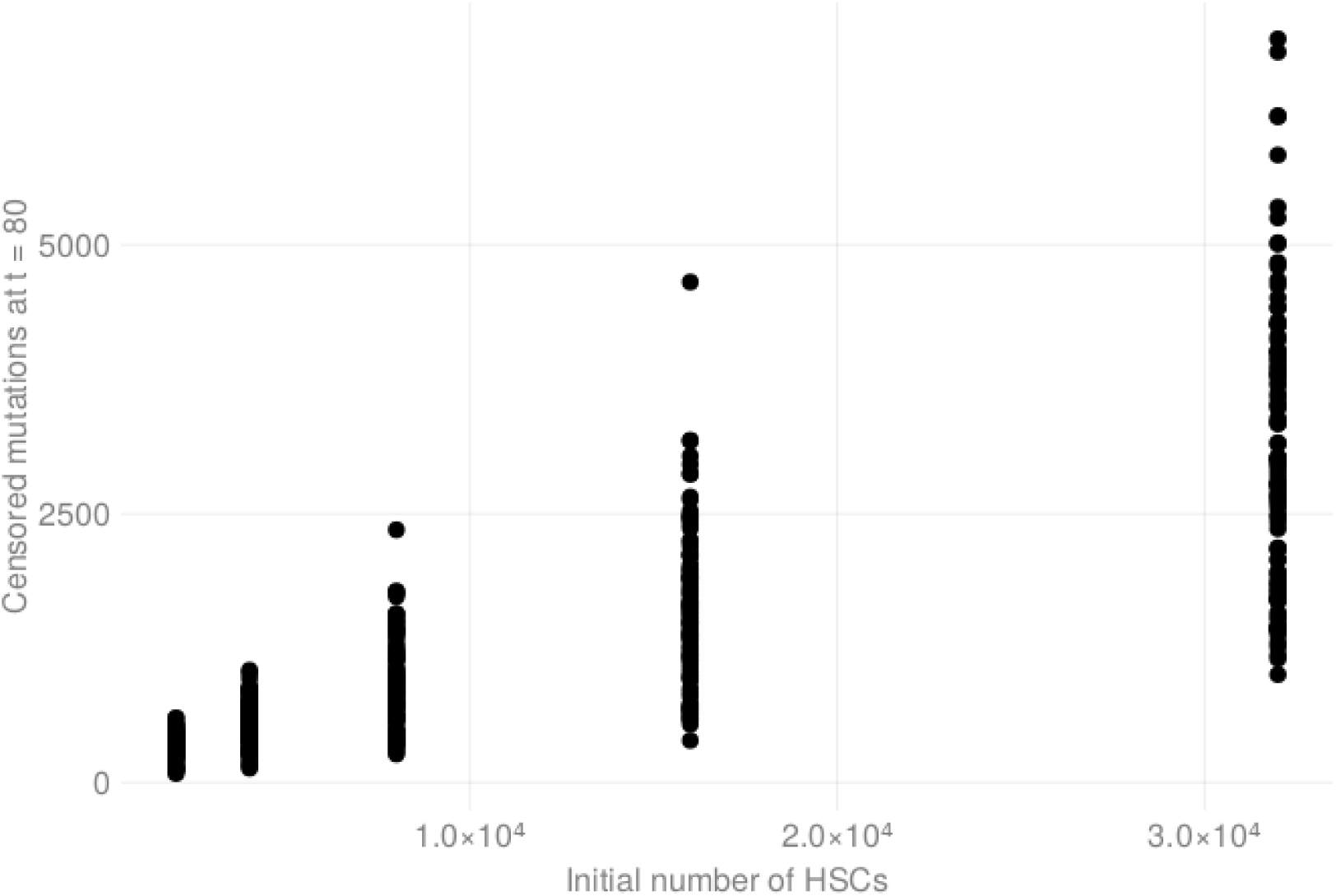
HSC stochastic process simulation, showing that the number of active HSCs has a large effect on the number of high-VAF mutations at the end of the simulation

**Extended Data Figure 5:**
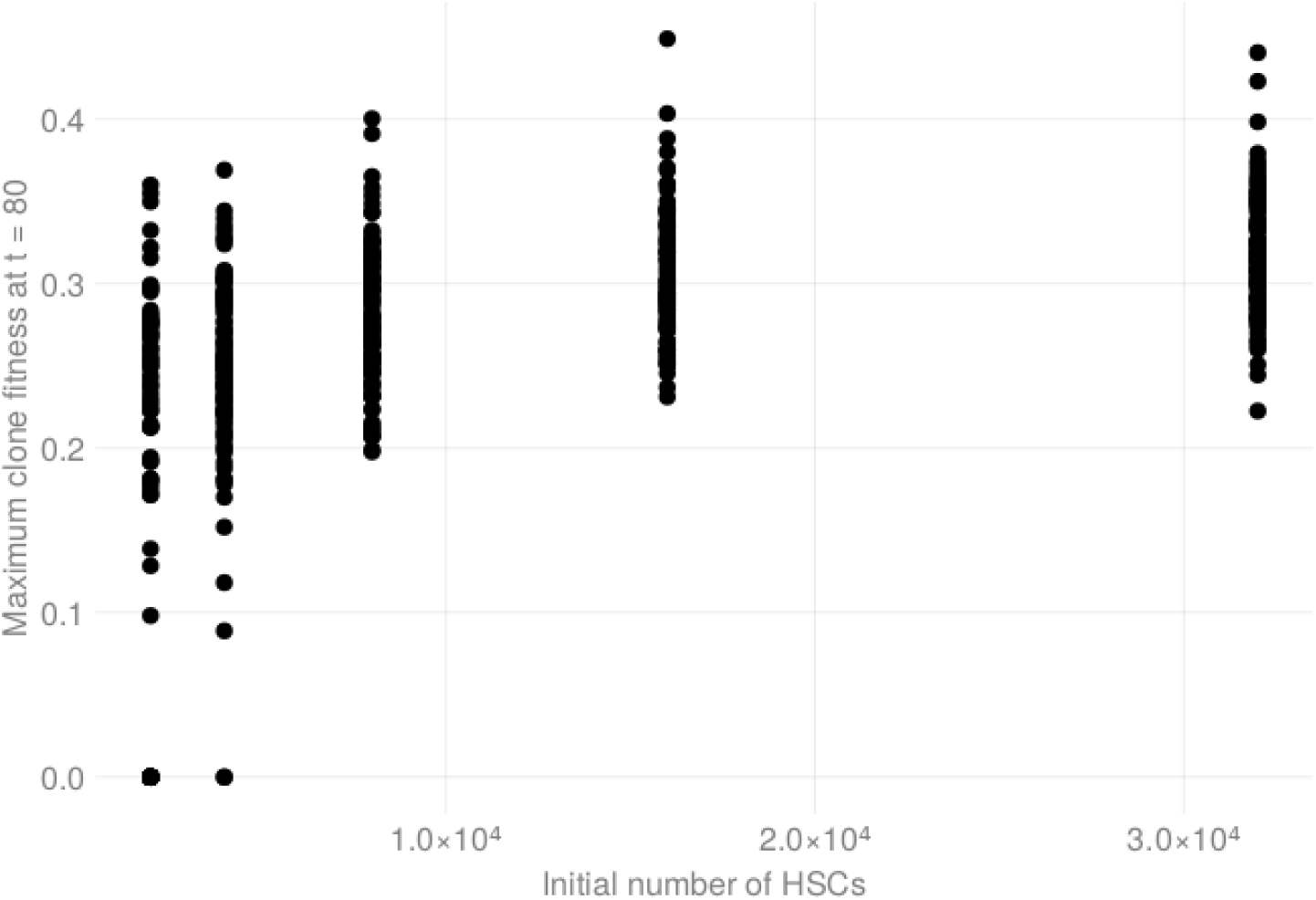
HSC stochastic process simulation, showing that the number of active HSCs has a large effect on likelihood of obtaining at clone with high fitness

**Extended Data Figure 6:**
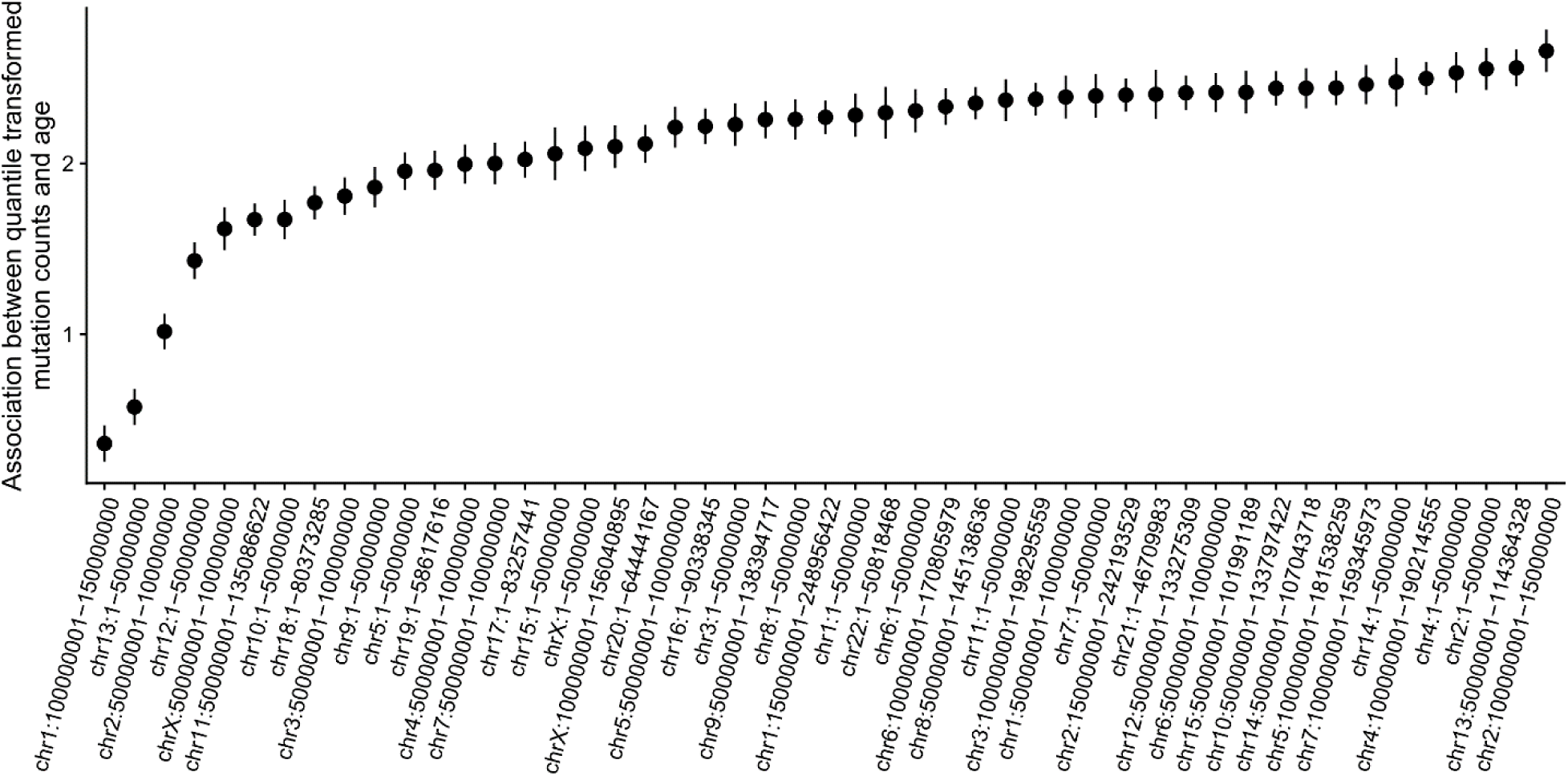
Linear regressions were performed between the inverse normal transformed mutation burden in each genomic bin with chronological age on the y-axis. Each regression include a study indicator as a covariate.

**Extended Data Figure 8:**
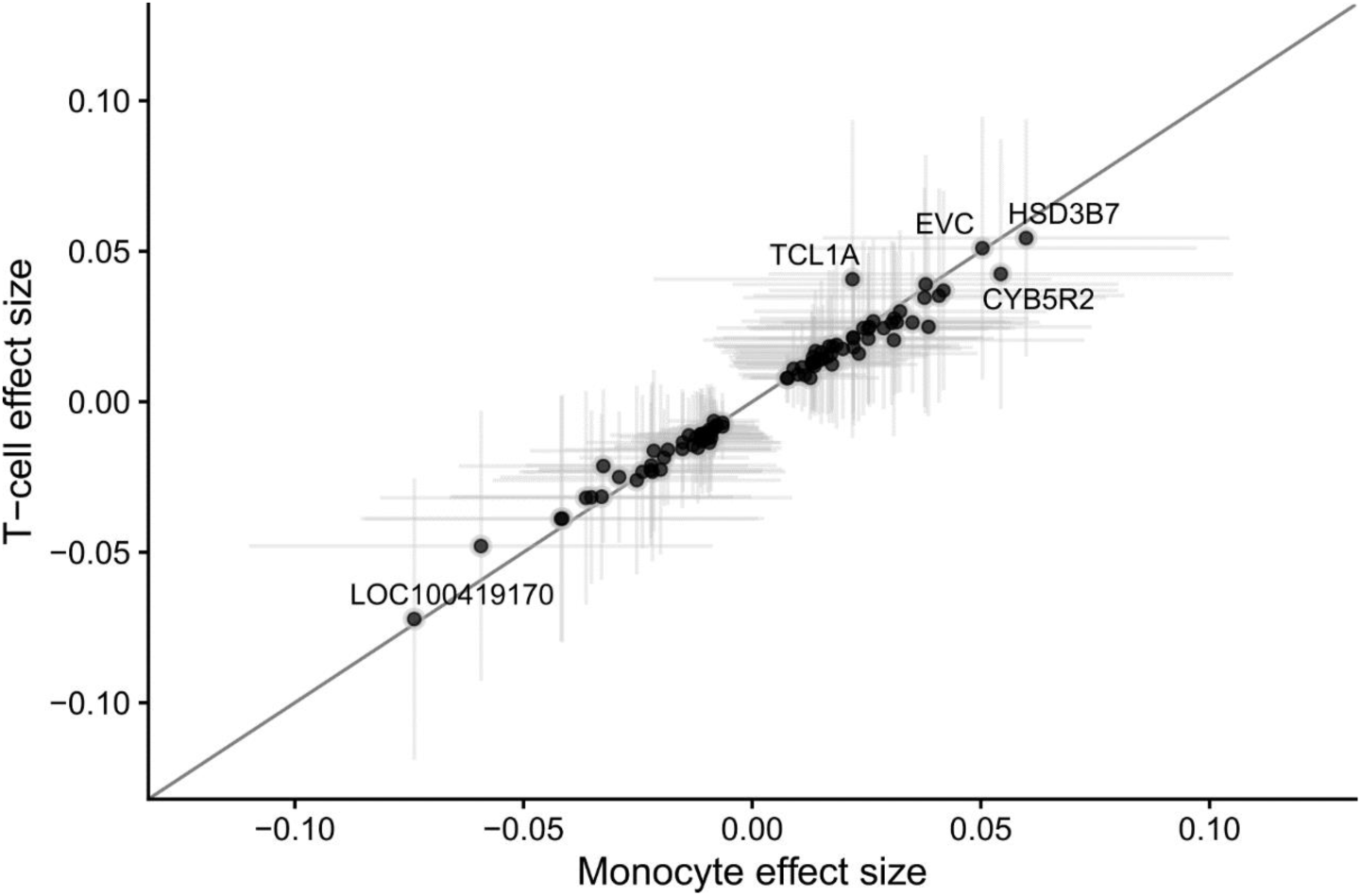
The association between GEM and gene expression in either monocytes or T cells. Effect sizes are estimated after application of mashr shrinkage, and the intervals denote 95% credible intervals.

**Extended Data Figure 8:**
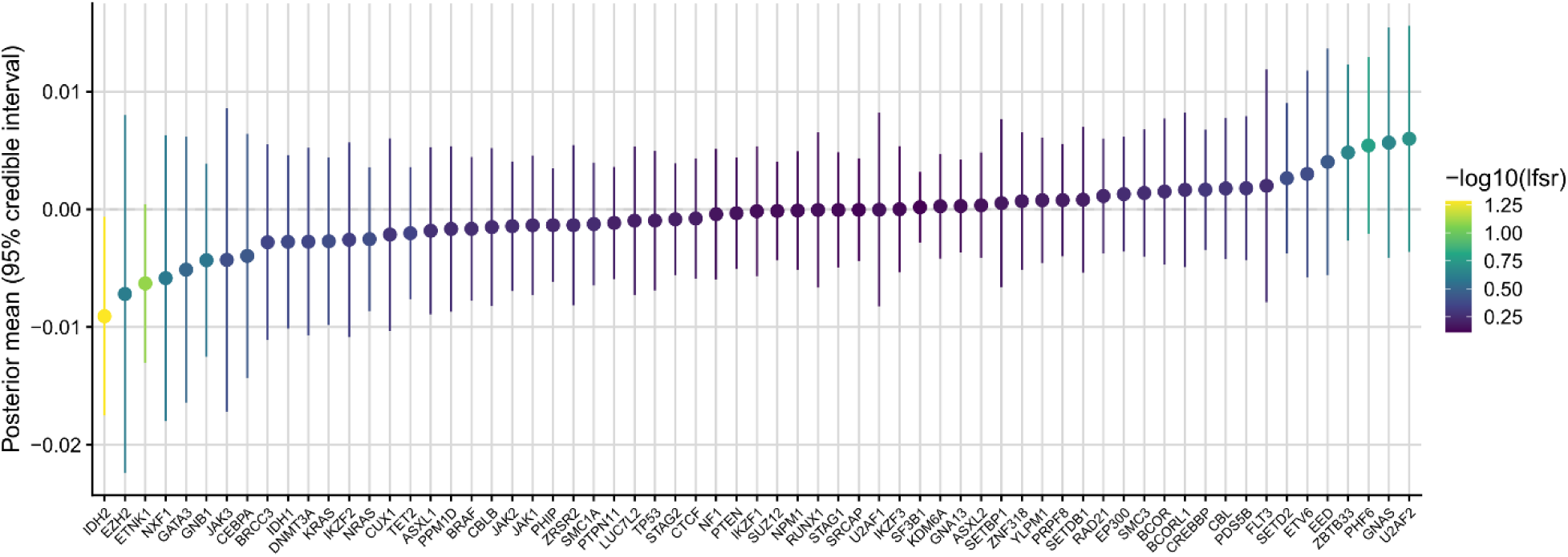
The association between GEM and gene expression in whole blood among CHIP genes. Effect sizes are estimated after application of mashr shrinkage, and the intervals denote 95% credible intervals.

**Extended Data Figure 9:**
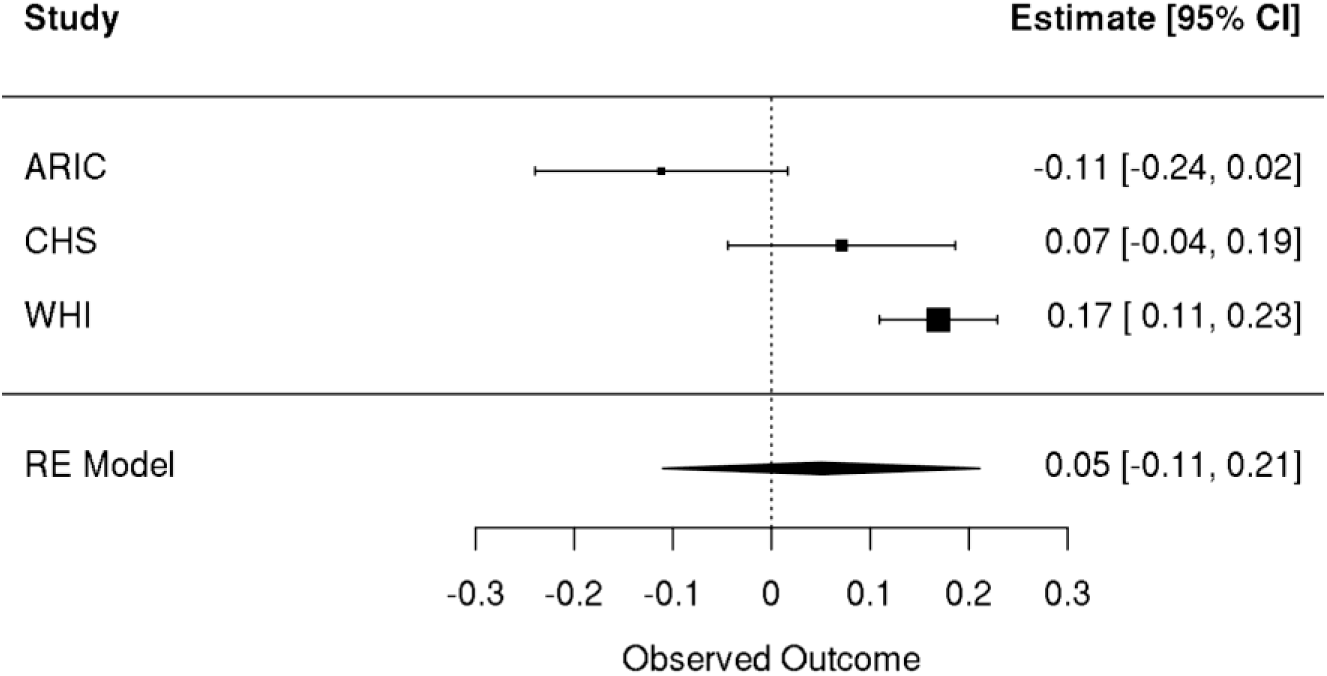
Meta-analyses of Cox proportional hazards regression with time to ischemic stroke as the outcome. A spline of age, sex, smoking status, and germline PCs were included as covariates. Individuals with prevalent disease were excluded.

**Extended Data Figure 10:**
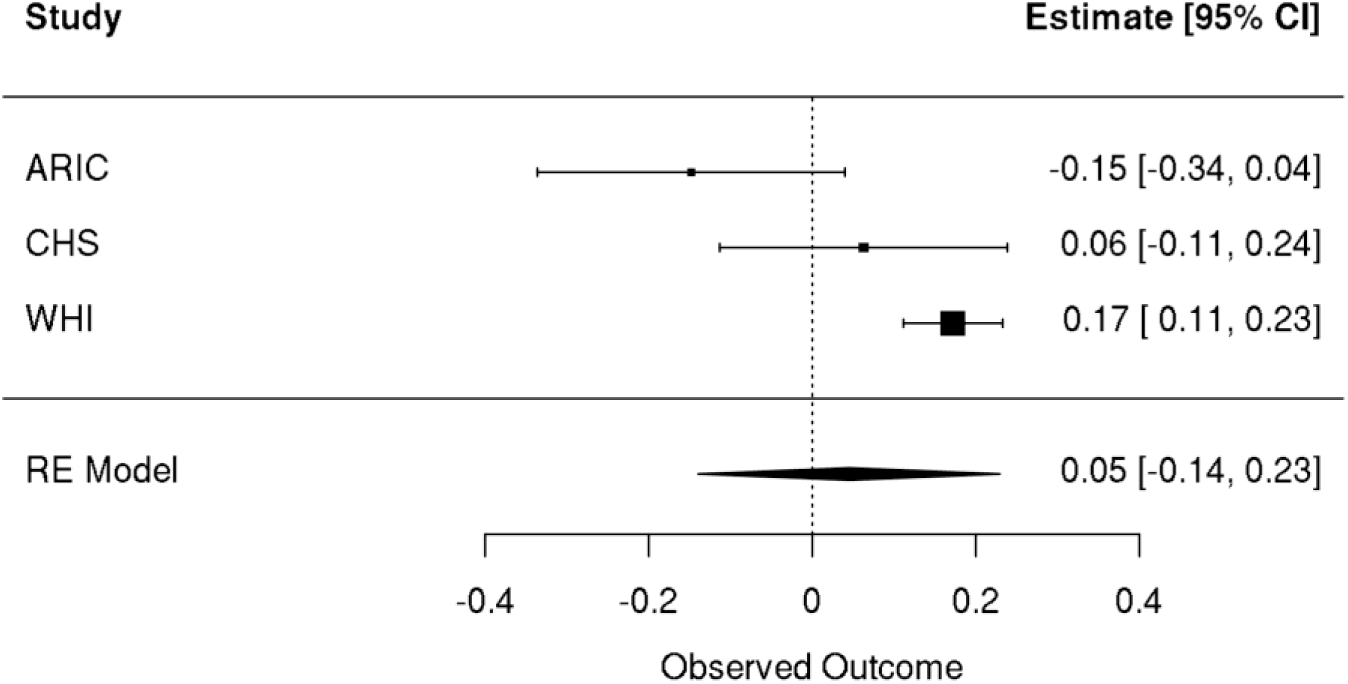
Female only meta-analyses of Cox proportional hazards regression with time to ischemic stroke as the outcome. A spline of age, sex, smoking status, and germline PCs were included as covariates. Individuals with prevalent disease were excluded.

**Extended Data Figure 11:**
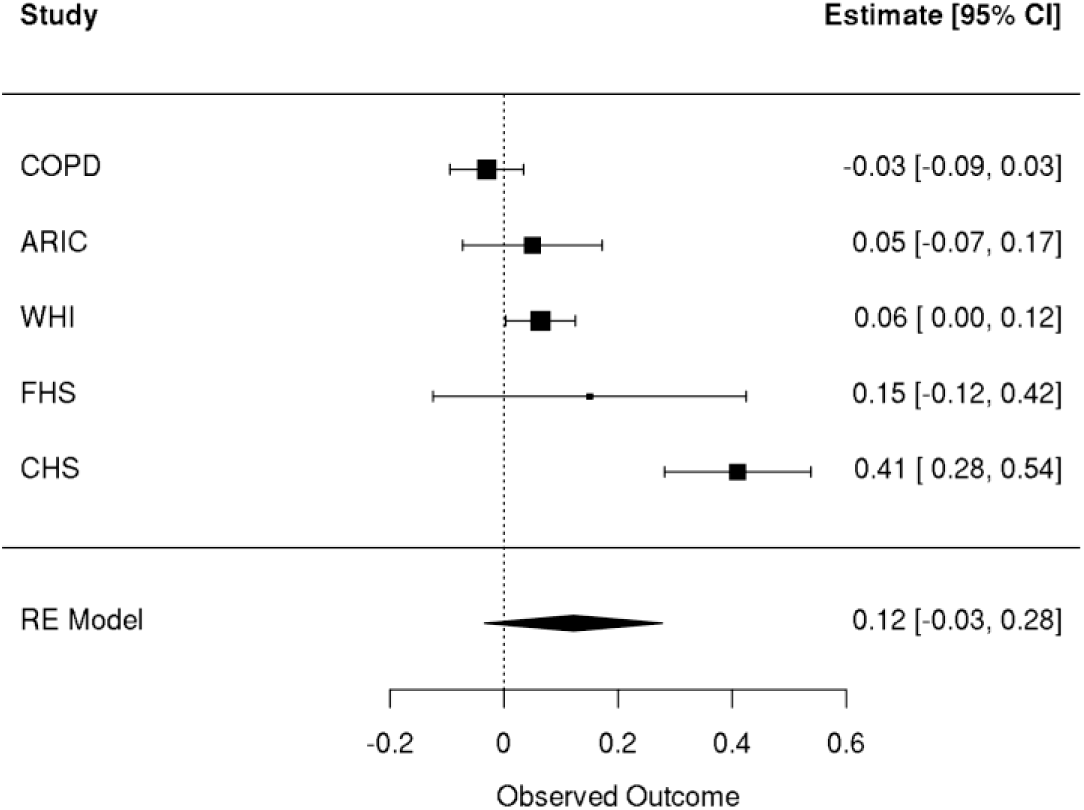
Meta-analyses of linear regressions with inverse normal transformed GEM as the outcome and an indicator for prevalent coronary artery disease events that occurred prior to the blood draw that GEM uses as the covariate of interest. A spline of age, sex, smoking status, and germline PCs were included as covariates.

## Methods

### Germline and somatic variant calling

TOPMed germline variant was performed as previously described^25^. Briefly, TOPMed BAM files were harmonized through the functionally equivalent pipeline^72^. Joint calling of germline SNPs and indels was performed with the Got-Cloud pipeline^73^. Samples were aligned to GRCh38. TOPMed germline SNP and indel freeze 10 was used in this analysis.

Putative somatic variants were first called with GATK Mutect2^74^ in “tumor-only” mode with largely default settings. A “panel of normals” was included to exclude sequencing artifacts. Variant calling was performed on Google Cloud using Cromwell^75^. Only bi-allelic variants that passed Mutect2 filters were included in downstream analyses. CHIP calling was performed as previously described^13,17^; briefly, the Mutect2 output was cross-referencing with a list of predominately loss-of-function and missense mutations in a curated set of genes^4,76^.

We first identified somatic mutations that occurred once across all individuals, as singleton passenger mutations have a stronger association with chronological age than non-CHIP recurrent somatic mutations^77^. On mutations on the X-chromosome, we halved the variant allele-fraction for all mutations. We then excluded several mutations based on the following filters:

1. All mutations with a depth less than 25x or greater than 100x
2. All mutations falling within low complexity sequence regions
3. All mutations in segmental duplications
4. All mutations falling within genomic regions with germline CNVs with at least 10% minor-allele frequency. Germline CNVs from the TOPMed germline structural variant call-set^78^ were used in this filter.
5. All mutations falling within the contigs with sequence that differed between hg19 and hg38, as defined by the “Hg19 diff” track in the UCSC genome table browser.
6. Any germline variant in TOPMed germline SNP and index freeze 10 (derived from 184,878 WGS) with a minor allele count of at least 10 and a variant allele fraction between .26 and .74
7. Any mutation with fewer than 2 alt reads or greater than 6 alt reads. At 38x, this corresponds to a VAF interval of 5%-16%

### Annotation of somatic mutations

Singleton mutations after the above filters were first annotated with the variant effect predictor^79^ (VEP) including the “—flag-pick”, “—check_existing”, “—canonical”, and “—flag_pick” flags. A CADD^33^ plugin was also included. CD34+ chromatin annotations were downloaded for sample BSS00233 from Roadmap epigenomics^29^. Mutations were also annotated with the correspond mutation type (e.g., C->T, G-T, etc.).

### The genomic and epigenomic mutation rate (GEM)

GEM is a Bayesian graphical model with the following form. In the outlier layer, standardized chronological age (standardized with (age – 60) / 10) is the outcome variable, denoted as *Y*_*i*_. We note that GEM is a “weakly-supervised” model in the sense that while individual mutations are unlabeled, the entire training process is “supervised” by chronological age. *Y*_*i*_ conditional on the number of true mutations within an individual is assumed to follow a gaussian distribution. Each individual *i* has a candidate set of mutations *S*_*i*_ which were identified by the above filtering processes. Instead of using this raw count, we instead replace the count with the expectation of a Bernoulli random variable *Z*_*i*,*j*_which denotes whether the *jt*ℎ mutation in the *it*ℎ individual is a “true mutation” (i.e., takes a value of 1.0) or is an artifact (i.e., takes a value of 0). We include a non-linear transformation *g* to the sum over the true mutation burden. In practice, *g*(*x*) = *log*2(*x*) worked well.

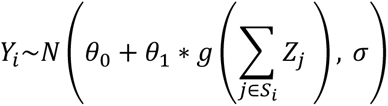

The expectation of this random variable is specified through an inverse-logit transformation, i.e.,

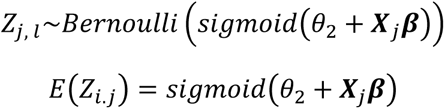

Where 𝑿_𝒋_ represents a length 𝑝 vector of annotations for the *jt*ℎ mutation and 𝜷 is a length 𝑝 random vector of weights. *θ*_2_is included as a bias or intercept term.

The above assumptions express the likelihood of GEM. The prior of GEM is specified as follows:

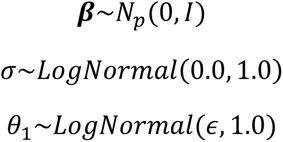

Where 𝜖 is in practice set to 5 x 10^-3^. Inference was performed by optimizing the maximum a-posteriori objective using an ADAM optimizer. GEM is implemented in the torch package in R.

Within the matrix of mutation annotations 𝑿, we include the following annotations:

1. VEP annotated variant impact
2. VEP “somatic” annotation
3. CADD_PHRED score
4. The mutation type
5. The variant allele fraction
6. The chromatin state prediction

GEM was trained on 2,000 randomly sampled individuals with 186,277 total candidate mutations among them.

### Genome-wide association studies with GEM

In the context of genome-wide association studies (GWAS), the phenotype was defined as the expected burden of “true” mutations, i.e., ∑_*j*∈*Si*_ *E*(*Z*_*j*_) within the *it*ℎ individual. This phenotype was inverse normal transformed. GWAS summary statistics were estimated with SAIGE on all germline variants where the minor allele count was at least 400 (i.e., MAF ≥ 0.4%) among the analyzed samples. Germline principal components 1-10, somatic principal components 3-4, genotype inferred sex, a cohort indicator, chronological age, average sequencing depth per sample, and the residual between the raw and estimated true mutation burden were included as covariates. Somatic principal components 1-2 were excluded as they are strongly associated with total mutation burden and sex respectively. Somatic mutations that had previously been identified as recurrent^77^ were excluded from the summary statistics. All germline variants with a milk-SVM threshold below -0.30 or an individual specific Hardy-Weinberg equilibrium -log10 pvalue above 5.0 were excluded.

### Rare-variant association studies with GEM

Rare-variant association studies (RVAS) were performed with the same GEM derived phenotype as the GWAS. We performed a non-coding RVAS by examining rare-variants within 100kb of cancer associated genes as defined by Open Targets^37^ using SCANG^56^, which performs a scanning procedure for genome regions that contain a set of rare variants that associate with the phenotype. We similarly performed a genome-wide coding variant RVAS using STAAR, including any rare-variant annotated as having a “MODERATE” or “HIGH” impact on amino acid sequence by VEP.

### Simulation of mutation burden

We assume an HSC can fall into one of three states:

1. HSCs can divide into two HSCs (“self-renewal”)
2. HSCs can divide into two differentiated cells
3. HSCs can divide into one HSC and one differentiated cell

For the purposes of simulating a stochastic process of HSC population size, we treat state 3 as irrelevant because it does not affect the total number of self-renewing HSCs.

We define the HSC clone birth rate as: 𝜆_*i*_(*t*) ∼ 𝑃𝑜*iss*𝑜𝑛(𝜔 ∗ 𝑋_*i*_(*t*) ∗ (1 + *s*_*i*_(*t*)) ∗ 𝑑*t*) and the HSC clone death rate as 𝜓_*i*_(*t*) ∼ 𝑃𝑜*iss*𝑜𝑛(𝜔 ∗ 𝑋_*i*_(*t*) ∗ (1 − *s*_*i*_(*t*)) ∗ 𝑑*t*). 𝜔 is a parameter that controls the rate of births/deaths. 𝑑*t* defines the time interval over which this process is defined.

The total size of the cells within the *it*ℎ clone at time *t* as 𝑋_*i*_(*t*) = ∑_𝑙≤*t*_ 𝜆_*i*_ (𝑙) − 𝜓_*i*_(𝑙). A single parameter *s*_*i*_ determines the likelihood of a given HSC falling into state 1. or 2., and thus we refer to this parameter as the clone “fitness.”

At any given time *t*, the VAF of the *it*ℎ clone is defined as 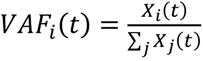. We define the number of passenger mutations at time *t* in the *it*ℎ clone as 𝐴_*i*_(*t*) ∼ 𝑃𝑜*iss*𝑜𝑛(𝑋_*i*_(*t*) ∗ 𝜇_𝑝_ ∗ 𝑑*t*), where 𝜇_𝑝_ is a per-cell passenger mutation rate. We define 𝐴𝐶_*i*_(*t*) as the count of “censored” passenger mutations at time *t* for the *it*ℎ clone, where the censoring occurs due to the limited sensitivity of ∼38x sequencing coverage. This censoring is implemented by the following probability 𝑃(𝐵*i*𝑛𝑜𝑚*i*𝑎𝑙(38, 𝑉𝐴𝐹_*i*_) > 2).

### Association between GEM and gene expression

Separate association analyses were performed for each of the five tissue types available within TOPMed: whole blood, PBMCs, T cells, monocytes, and nasal epithelial tissue. We performed linear regression between the inverse normalized GEM estimate of the true mutation burden and inverse normalized gene expression in the tissue, where chronological age, sex, germline genotype PCs 1-15 and expression PCs 1-20 were included as covariates. In the whole blood analysis, we also included a cohort indicator as a covariate. Summary statistics from each analysis were then included a Bayesian multivariate analysis implemented in mashr ^62^. As a measure of “significance”, we used the mashr estimate of the local false-sign rate (LFSR) < 0.05. Enrichment analyses were performed with the pathfindR^80^ package including all tested genes as the background set and Reactome^81^ as the reference database for gene sets.

### Incident ischemic stroke analysis

Ischemic stroke at most recent visit was chosen for the survival analysis event, and the time to event was defined as the difference in years between baseline and the most recent visit. The WHI, CHS, and ARIC cohorts were included. There was a total of 9,885 individuals included in this analysis from the WHI cohort. In WHI, 9,520 samples were included, there were 1,134 events. In CHS, 2,822 samples were included, and 199 had events. For ARIC, 3,475 samples were included with 231 events. Covariates included Ischemic case status at baseline, BMI measured at baseline, "ever smoker" status at baseline, a spline of age at blood draw, and genetic ancestry PCs 1-4.

### Incident coronary artery disease analysis

Incident coronary artery disease at most recent visit was chosen for the survival analysis event, and the time to event was defined as the difference in years between baseline and the most recent visit. A composite coronary artery disease phenotype was defined as an event if at least one of the following occurred during the follow-up period: myocardial infarction, coronary artery bypass graft, angina, angioplasty, or death due to coronary heart disease. Individuals with prevalent disease based on this composite phenotype were excluded. The WHI, CHS, COPDGene, and FHS cohorts were included. In WHI, 9,039 samples were included, there were 1,787 events. In CHS, 2,456 samples were included, and 933 had events. In FHS, 3,786 samples were included and 525 had events. For COPD, 4,987 samples were included with 133 events. Covariates included Ischemic case status at baseline, BMI measured at baseline, "ever smoker" status at baseline, a spline of age at blood draw, and genetic ancestry PCs 1-4.

### Lentiviral transduction of healthy CD34+ cells

Lentiviral vectors expressing NRIP1 (V2LHS_172503, V2LHS_172504 and V2LHS_172507) or SMC4-targeting shRNA (V2LHS_21882, V3LHS318029, V3LHS_318030) (Horizon) or non-silencing pGIPZ-puro lentiviral vector was transfected together with pCMV-dR8.9 and vesicular stomatitis virus G–expressing plasmids into HEK 293-FT cells using Lipofectamine 2000 (Thermo Fisher Scientific) for lentiviral supernatant production as previously described^82^. Primary CD34+ cells were obtained as excess material from harvests of normal donors for allogeneic bone marrow transplantation. Specimens were collected by the Johns Hopkins Kimmel Cancer Center Specimen Accessioning Core. Appropriate informed consent was obtained from all donors before specimen collection in accordance with the Declaration of Helsinki and under a research protocol approved by the Johns Hopkins Institutional Review Board. CD34+ cell subsets were isolated using the CD34 MicroBead kit (Miltenyi Biotec) as previously described^83^. CD34+ cells were incubated with the viral supernatant and polybrene (8µg/ml; MilliporeSigma) for transduction in wells pre-coated with retronectin (20ng/ml; MilliporeSigma). After at least 48 hours, cells were treated with puromycin (0.5µg/ml; MilliporeSigma) for 4 days to select resistant cells.

### Apoptosis and differentiation assays

Apoptosis was assessed by 7-AAD staining evaluated by flow cytometry (Thermo Fisher Scientific #00-6993-50). Percentages of stem (CD34+CD38-) and progenitor cells (CD34+CD38+) were assessed by CD34 (Thermo Fisher Scientific #11-0349-42) and CD38 (BioLegend #356641) staining evaluated by flow cytometry.

### Clonogenicity assays

CD34+ cells following puromycin treatment were collected, counted, and plated at a density of 2000 cells/ml in methylcellulose-based media as previously described^82^. After 10 to 14 days of incubation at 37°C in 5% CO2, the recovery of colony-forming units (burst forming unit-erythroid (BFU-E) and colony forming unit-granulocyte/monocyte (CFU-GM)) were determined by colony counting under bright-field microscopy. A cell aggregate composed of >50 cells was defined as a colony.

## Supporting information

Supplementary cohort acknowledgements

## Data Availability

Individual whole-genome sequence data for TOPMed whole genomes, individual-level harmonized phenotypes and the CHIP variant call sets used in this analysis are available through restricted access via the dbGaP TOPMed Exchange Area available to TOPMed investigators.

## Code and data availability

Code for the Genomic and Epigenomic Mutation rate pipeline: https://github.com/weinstockj/GEM. Individual whole-genome sequence data for TOPMed whole genomes, individual-level harmonized phenotypes and the CHIP variant call sets used in this analysis are available through restricted access via the dbGaP TOPMed Exchange Area available to TOPMed investigators.

## Acknowledgements

Whole genome sequencing (WGS) for the Trans-Omics in Precision Medicine (TOPMed) program was supported by the National Heart, Lung and Blood Institute (NHLBI). See Supplementary Information 1 for study omics support information. Centralized read mapping and genotype calling, along with variant quality metrics and filtering were provided by the TOPMed Informatics Research Center (3R01HL-117626-02S1; contract HHSN268201800002I). Phenotype harmonization, data management, sample-identity quality control and general study coordination were provided by the TOPMed Data Coordinating Center (R01HL-120393; U01HL-120393; contract HHSN268201800001I). We thank the studies and participants who provided biological samples and data for TOPMed. The full study-specific acknowledgments are included in Supplementary Cohort Acknowledgements. The views expressed in this manuscript are those of the authors and do not necessarily represent the views of the National Heart, Lung, and Blood Institute; the National Institutes of Health; or the US Department of Health and Human Services. The authors wish to acknowledge the contributions of the consortium working on the development of the NHLBI BioData Catalyst ecosystem.

